# Subthalamic DBS Engages Right-lateralized Frontal Control to Improve Gait Adaptation in Parkinson’s

**DOI:** 10.64898/2026.06.03.26354536

**Authors:** Ibrahem Hanafi, Nicolo G. Pozzi, Rita Habib, Salvatore Falciglia, Jasmin Del Vecchio Del Vecchio, Luigi Gianmaria Remore, Giorgio Marotta, Andreas Buck, Gianni Pezzoli, Jens Volkmann, Ioannis U. Isaias, Chiara Palmisano

## Abstract

Adapting ongoing gait patterns to environmental challenges is essential for safe navigation through the environment. Impairment of gait adaptation is common in many neurodegenerative disorders, such as Parkinson’s disease (PD), where it hampers mobility and limits quality of life. The neural control of gait adaptation remains largely unclear, thereby limiting the development of targeted treatments, such as deep brain stimulation of the subthalamic nucleus (STN-DBS). We integrated clinical, kinematic, brain metabolic imaging, and electrophysiological data, obtained during a fully immersive virtual reality overground walking task, to characterize the neural underpinnings of gait adaptation performance during dynamic obstacle avoidance and its improvement with STN-DBS.

Movement kinematics, brain oscillatory activity, and metabolic activation were simultaneously acquired in 12 patients with PD during rest and gait adaptation, under active or paused STN-DBS, using inertial measurement units, electroencephalography, and three separate [18F]fluorodeoxyglucose positron emission tomography scans. Eight age-matched healthy subjects completed the same task for comparative kinematic analyses.

All patients showed significant clinical improvement with STN-DBS. During the gait adaptation task with paused stimulation, patients exhibited increased metabolic activity in the cerebellum and sensorimotor cortex. Active STN-DBS selectively enhanced thalamic and superior frontal gyrus (SFG) metabolism, while concomitantly reducing cerebellar uptake. Right-lateralized SFG metabolism correlated with gait adaptation performance, with DBS-driven shifts toward greater right SFG activity predicting the magnitude of gait adaptation improvement. This correlation was independent of baseline asymmetry in clinical impairment, electrode placement, or structural connectivity to the SFG. Of note, STN-DBS amplitude asymmetry emerged as an independent predictor of right-lateralization of SFG metabolism. EEG recordings confirmed this lateralized network modulation, with theta-band asymmetry paralleling PET findings.

Our findings identify a lateralized thalamo-cortical network supporting gait adaptation in PD and highlight a distinctive role for the SFG. We further show that effective STN-DBS acts as a lateralized regulator, dynamically rebalancing cortico-thalamic circuits to support context-appropriate gait control. The observed right-hemispheric lateralization may foster novel image-guided programming strategies to enhance the consistency and effectiveness of gait control in PD.

## 2. Introduction

Gait is a fundamental motor behavior enabling effective interaction with the environment. The safe execution of walking critically depends on the capacity to dynamically adapt locomotor patterns to environmental demands.^1–5^ Impairment of gait adaptation is a common feature of several neurodegenerative disorders, including Parkinson’s disease (PD),^6,7^ where it represents a major cause of disability and reduced quality of life.^8–10^

Deep brain stimulation of the subthalamic nucleus (STN-DBS) is an established treatment that significantly improves motor symptoms in PD.^11^ However, its effects on gait and postural control remain incompletely characterized,^3^ with some patients experiencing limited benefit or even deterioration.^12,13^ Notably, only a limited number of studies have specifically investigated the effects of STN-DBS on gait adaptability.^14–17^ Existing work has largely relied on simplified or highly artificial experimental paradigms, such as treadmill walking, stepping tasks, or discrete gait transitions, which may not fully capture the complexity of real-world locomotion.^6,18–20^ As a result, how STN-DBS modulates the neural processes underlying adaptive gait in ecologically valid conditions remains poorly understood, leaving critical aspects of its network-level mechanisms unresolved.^3^

Gait adaptation relies on a distributed brain network that supports executive control, sensorimotor integration, and cognitive-limbic modulation within a hierarchical locomotor system involving frontoparietal, basal ganglia, cerebellar, and brainstem circuits.^21–23^ Within this framework, the frontal lobe, particularly the superior frontal gyrus (SFG), plays a key role in attention allocation, dual-task processing, and the goal-directed control of gait.^24,25^ Notably, dysfunction of the SFG has been associated with dopaminergic deficits in the caudate nucleus in PD,^26^ suggesting that impaired gait adaptation in PD reflects a broader disruption of dynamic motor control involving frontal-basal ganglia networks.^27^

Emerging evidence further indicates a degree of hemispheric specialization within frontal locomotor networks, which may support the integration of spatial and executive demands during adaptive locomotion.^28,29^ Its disruption, in turn, has been associated with impaired gait automaticity and increased reliance on visual and cognitive control mechanisms, as observed in parkinsonian gait.^30–34^ In this context, hemispheric asymmetries in network engagement may contribute to variability in clinical responses to STN-DBS.^35,36^ Programming strategies based on a single hemisphere, often guided by acute motor responses, may fail to capture the distributed, long-range, and interhemispheric effects of STN-DBS,^37^ thereby potentially impairing large-scale network interactions underlying effective adaptive gait control.^6^

In this study, we combined clinical, kinematic, brain metabolic imaging, and electrophysiological data to investigate the neural underpinnings of gait adaptation impairments in PD. Using a fully immersive virtual reality (VR) paradigm that allows controlled yet ecologically relevant assessment of adaptive locomotion, we show that STN-DBS enhances gait control by inducing lateralized, behaviorally relevant changes in neural activity.

## 3. Materials and methods

### 3.1. Subjects and surgery

We investigated 12 right-handed subjects (three females) with idiopathic PD diagnosed according to the UK Brain Bank Criteria.^38^ Inclusion criteria were bilateral STN-DBS with stable stimulation settings and antiparkinsonian medications for at least six weeks prior to participation. Subjects with comorbid conditions potentially affecting gait or balance (e.g., vestibular or orthopedic disorders) or cognitive impairment were excluded. All participants underwent DBS implantation at the University Hospital of Würzburg; the surgical procedure has been described previously.^39,40^

Clinical assessments were performed on the first day of the neuroimaging acquisition under the same treatment conditions, namely the medication-off (meds-off) state (i.e., after overnight withdrawal of all antiparkinsonian medications) and either the stimulation-off (stim-off) condition (i.e., 1 h after DBS was switched off) or the stimulation-on (stim-on) condition (i.e. under chronic stimulation settings).

Disease stage was assessed using the Hoehn and Yahr scale (H&Y). The severity of motor impairment was evaluated using the MDS-UPDRS Part III. Sub-scores for bradykinesia, rigidity, tremor, and axial symptoms were derived from the MDS-UPDRS-III.^41^ To specifically capture postural instability and gait disorder (PIGD), we calculated an axial sub-score comprising items 3.10, 3.11, and 3.12.^42,43^ DBS-related clinical improvement was quantified as the absolute difference between stim-on and stim-off MDS-UPDRS-III scores (Supplementary table 1).

The most-affected hemibody was defined as the side with the higher summed score across MDS-UPDRS-III items assessing bradykinesia, rigidity and tremor, each rated separately for the left and right hemibody (i.e., from 3.3 to 3.8 and from 3.15 to 3.17).^44^ These same items were used to compute the asymmetry index of clinical impairment (AI Clinic) according to the following formula (Table 1):

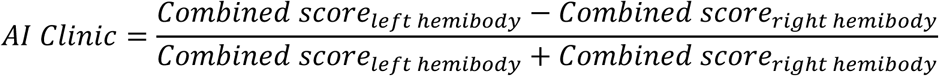

**Table 1:**
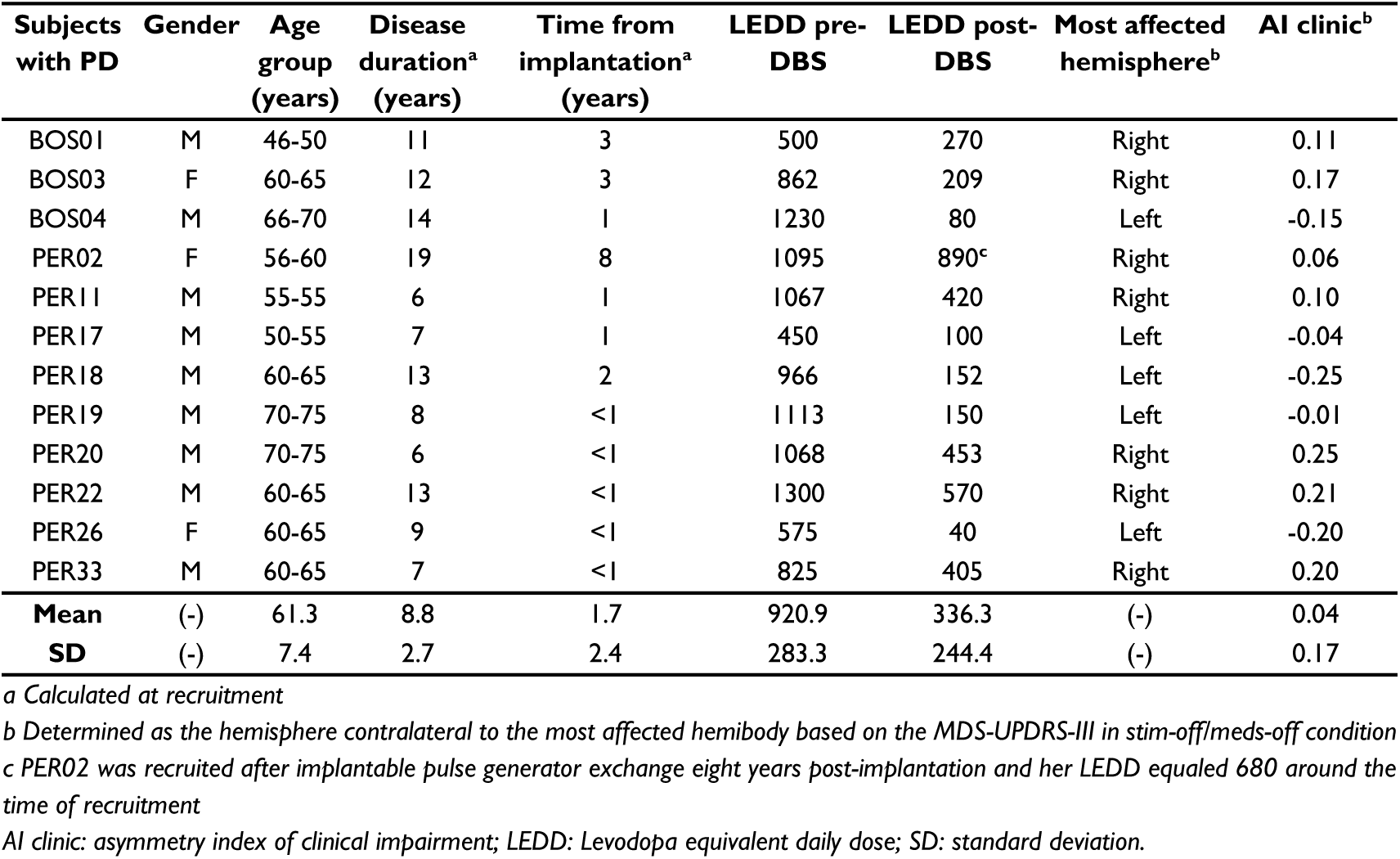
The demographic and clinical characteristics of subjects with PD

| Subjects with PD | Gender | Age group (years) | Disease duration<sup>a</sup> (years) | Time from implantation<sup>a</sup> (years) | LEDD pre-DBS | LEDD post-DBS | Most affected hemisphere<sup>b</sup> | AI clinic<sup>b</sup> |
| --- | --- | --- | --- | --- | --- | --- | --- | --- |
| BOS01 | M | 46-50 | 11 | 3 | 500 | 270 | Right | 0.11 |
| BOS03 | F | 60-65 | 12 | 3 | 862 | 209 | Right | 0.17 |
| BOS04 | M | 66-70 | 14 | 1 | 1230 | 80 | Left | -0.15 |
| PER02 | F | 56-60 | 19 | 8 | 1095 | 890<sup>c</sup> | Right | 0.06 |
| PER11 | M | 55-55 | 6 | 1 | 1067 | 420 | Right | 0.10 |
| PER17 | M | 50-55 | 7 | 1 | 450 | 100 | Left | -0.04 |
| PER18 | M | 60-65 | 13 | 2 | 966 | 152 | Left | -0.25 |
| PER19 | M | 70-75 | 8 | <1 | 1113 | 150 | Left | -0.01 |
| PER20 | M | 70-75 | 6 | <1 | 1068 | 453 | Right | 0.25 |
| PER22 | M | 60-65 | 13 | <1 | 1300 | 570 | Right | 0.21 |
| PER26 | F | 60-65 | 9 | <1 | 575 | 40 | Left | -0.20 |
| PER33 | M | 60-65 | 7 | <1 | 825 | 405 | Right | 0.20 |
| **Mean** | (-) | 61.3 | 8.8 | 1.7 | 920.9 | 336.3 | (-) | 0.04 |
| **SD** | (-) | 7.4 | 2.7 | 2.4 | 283.3 | 244.4 | (-) | 0.17 |

<sup>a</sup> Calculated at recruitment
<sup>b</sup> Determined as the hemisphere contralateral to the most affected hemibody based on the MDS-UPDRS-III in stim-off/meds-off condition
<sup>c</sup> PER02 was recruited after implantable pulse generator exchange eight years post-implantation and her LEDD equaled 680 around the time of recruitment
AI clinic: asymmetry index of clinical impairment; LEDD: Levodopa equivalent daily dose; SD: standard deviation.

The levodopa equivalent daily dose (LEDD) was calculated preoperatively and at 1-year postoperatively.^45^ Medications remained stable throughout the study period.

Eight subjects were implanted with the Percept PC device and SenSight leads (7/8 subjects) or model 3389 leads (1/8 subjects) (Medtronic, PLC, USA). The remaining three participants received a Genus R16 (2/3 subjects) or Gevia (1/3 subjects) device with Cartesia directional leads (Boston Scientific, Inc., USA).

We calculated an asymmetry index of stimulation current intensity (AI Stim) using the following formula (Supplementary table 2):

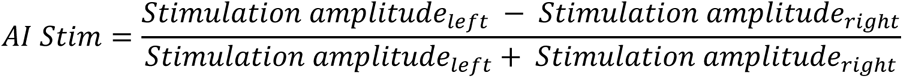

### 3.2. Study protocol and experimental setup

This study was approved by the Institutional Review Board of the University of Würzburg (No. 103/20-am) and conducted in accordance with the Declaration of Helsinki.^46^ Written informed consent was obtained from all participants.

We assessed brain metabolic activity using [18F]fluorodeoxyglucose positron emission tomography (FDG-PET) under three conditions: (i) resting state with meds-off/stim-off [Resting]; (ii) perturbed gait with meds-off/stim-off [Walk-off]; and (iii) perturbed gait with meds-off/stim-on [Walk-on]. Assessments were performed in the morning on three non-consecutive days after overnight fasting.

For the Resting and Walk-off scans, stimulation was switched off ≥30 min before FDG injection and remained off throughout the task and until completion of the scan. For the Walk-on scan, stimulation remained active throughout the experiment. Participants were continuously monitored by a neurologist (I.U.I. or N.G.P.), and none exhibited tremor or other symptoms that could interfere with FDG uptake.

Tasks (Resting or Walk) were performed for 10 min before FDG injection (preconditioning) and for at least 20 min during the 30-min FDG uptake period prior to scanning. This experimental design has previously been shown to identify gait-related brain metabolic activity.^47^

During the Resting condition, participants sat comfortably in a quiet room with eyes open, fixating a target positioned 5 m ahead.

For the Walk-off and Walk-on conditions, participants performed a gait adaptation task in a quiet room. A fully immersive VR environment was presented via a head-mounted display (Vive Pro, HTC). Participants walked barefoot overground at a self-selected speed, back and forth along a 10-m virtual walkway, while a virtual agent (VA) repeatedly crossed their path, requiring adaptive gait adjustments to avoid collisions. The VA motion was triggered by the participant entering a 3 m radius around the VA starting position, after which the VA moved at a speed equal to 1.5 times the participant’s instantaneous walking velocity recorded at VA movement onset. Further details of the VR environment have been reported previously.^48^ Eight age- and sex-matched healthy controls (two females; age 63.3 ± 5.8 years) performed the same walking task without undergoing PET imaging to provide a reference for gait performance.

Gait kinematics were recorded using three inertial measurement units (IMUs; sampling rate 128 Hz, Opal, APDM, USA) placed on the sternum (xiphoid process) and the lateral sides of the ankles. Head-mounted visor trajectories were analyzed to quantify gait adaptation performance. Cortical electrophysiological activity was recorded using a portable 64-channel electroencephalography system (EEG, sampling rate 500 Hz; Sessantaquattro, OT Bioelettronica, Italy).

The VR, IMU, and EEG recordings were synchronized using a transistor-transistor logic (TTL) signal, as previously described.^6^

### 3.3. Gait kinematic data processing

The VR system recorded the following: (i) the three-dimensional trajectory of the head-mounted visor; (ii) the distance between the participant and the VA over time (subject–VA distance, SVAD); (iii) the onset of the VA movement towards the participant (VA start); and (iv) the moment at which the VA crossed the participant’s path (VA cross).

To characterize the temporal window of gait adaptation, velocity profiles were computed as the first derivative of the visor trajectory along the walking direction (anterior–posterior axis). The resulting signals were low-pass filtered at 5 Hz (3rd-order Butterworth) and used to identify three reference points: (i) the start of the gait adaptation window (AW); (ii) the time of minimum velocity; and (iii) the end of the AW.

For each trial, minimum velocity was defined as the global minimum of the velocity profile. A steady-state velocity (SSV) range was computed as the mean ±1 standard deviation of velocity during the 1 s period preceding VA start. AW onset was defined as the last velocity peak preceding the minimum velocity, provided that velocity remained above the lower bound of the SSV range for at least 0.5 s. AW end was defined as the first velocity peak following the minimum velocity, provided that velocity remained above the lower bound of the SSV range for at least 0.5 s (Figure 1).

**Figure 1:**
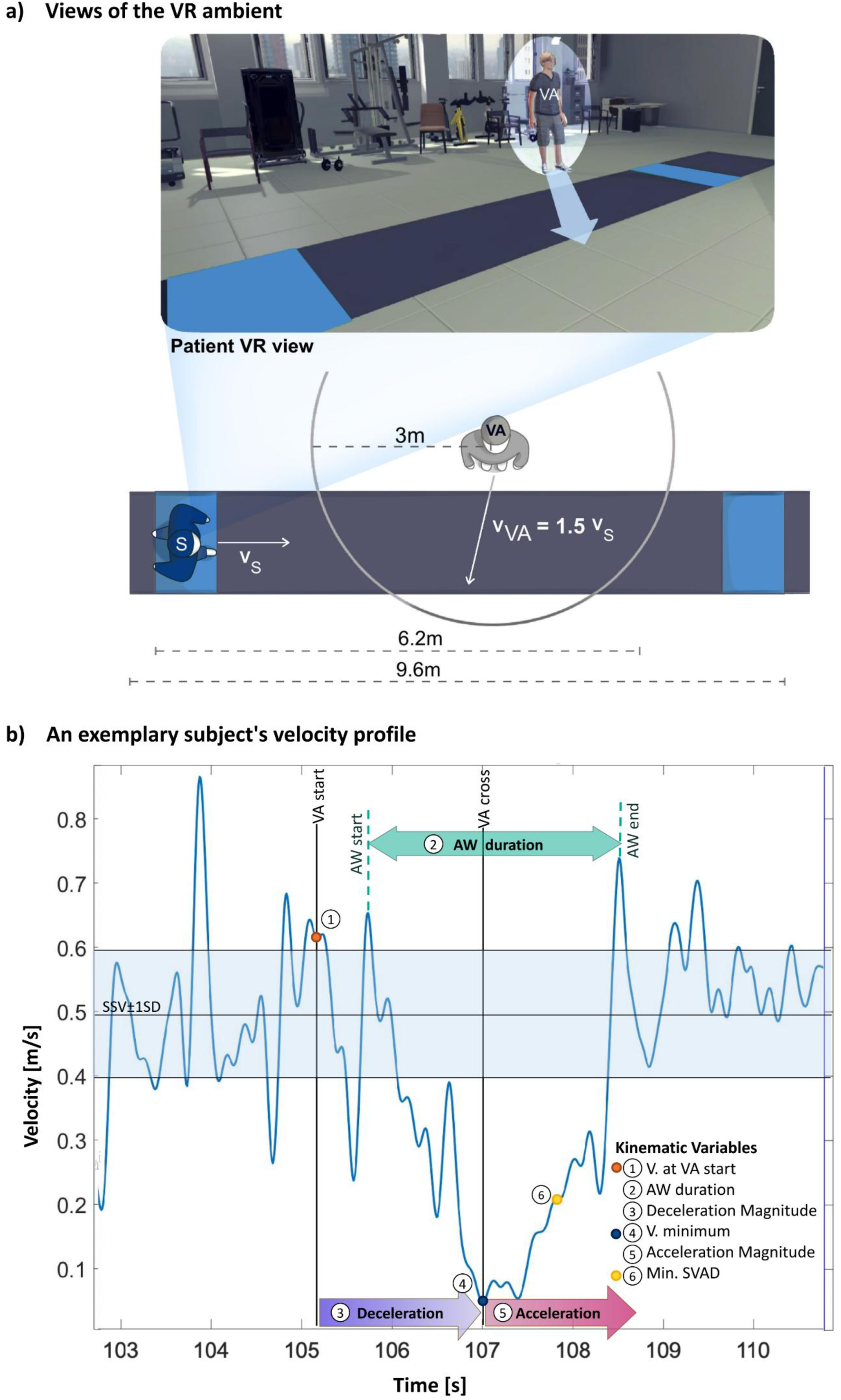
Views of the VR paradigm and an exemplary velocity profile of one subject during the task. a) A patient’s view of the VR ambient and a top-view scheme of the VR paradigm with the relative positions of the subject (S) and the VA at the start of the trial. b) An exemplary velocity profile (blue line) and kinematic variables of interest. The black horizontal line and the light blue area represent the mean SSV and the SSV range (mean ± 1SD), respectively, computed in the last second before the VA starts. Abbreviations: AW: adaptation window; SSV: steady-state velocity; SVAD: subject-VA distance; V.: velocity; VA: virtual agent.

Using these reference points, the following kinematic variables were derived to characterize gait adaptation: (i) velocity at VA start, representing the approach velocity to the obstacle; (ii) AW duration, defined as the interval between AW start and AW end; (iii) average deceleration, computed as the ratio between velocity change and elapsed time from VA start to minimum velocity; (iv) minimum velocity, defined as the lowest velocity reached during the AW; (v) average acceleration, computed as the ratio between velocity change and elapsed time from minimum velocity to AW end; (vi) minimum SVAD, defined as the minimum subject-VA distance, representing obstacle clearance.

In addition, 180° turning events at the ends of the walkway were detected from peaks in sternum angular velocity -low-pass filtered at 1.5 Hz with 3rd-order Butterworth filter to reduce high-frequency noise-around the vertical axis, reflecting shoulder-girdle rotation, and turn peak angular velocity was extracted.^49–51^

All kinematic analyses were performed offline in MATLAB R2022b (MathWorks, Inc., Natick).

Only trials occurring between FDG injection and PET acquisition were included in the kinematic analysis. Trials were excluded based on visual inspection (0–5 trials excluded per subject) if the task was interrupted, or gait adaptation was not evident.

For each participant, an equal number of trials from Walk-off and Walk-on conditions was analyzed; when necessary, the condition with more trials was truncated by selecting the first consecutive number of trials matching the condition with fewer trials. One participant (PER26) was excluded from the whole kinematic analysis, because of a technical recording failure, and two participants (PER19 and PER20) were excluded from the turning analysis only because they could not perform sufficient unaided turns.

To reduce dimensionality, the six kinematic variables listed above were subjected to principal component analysis (PCA) separately for the Walk-off and Walk-on conditions. Varimax rotation with Kaiser normalization was applied, and variables with absolute loadings <0.70 were suppressed. A composite gait-adaptation score was computed for each condition by averaging the Z-scores of kinematic variables contributing to the significant principal components across both conditions. Z-scores were computed after transforming variables such that higher (positive) values indicated better performance (i.e., closer to the HC range).^52^

To ensure comparability between stimulation conditions, and to account for potential differences in variance, Walk-on composite gait-adaptation scores were rescaled to the Walk-off distribution using min–max transforms:

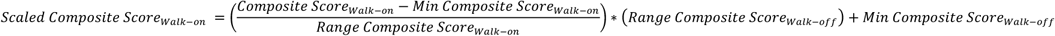

The DBS-related improvement in gait adaptation was defined as the difference between the scaled Walk-on composite score and the baseline Walk-off composite score:

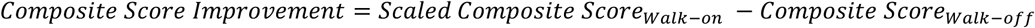

This measure reflects changes in each participant’s relative ranking under STN-DBS rather than absolute changes in kinematic performance. Similarly, Z-scores of SSV and turn peak angular velocity were computed for Walk-off and Walk-on, followed by min–max scaling and calculation of DBS-related change using the same procedure to estimate STN-DBS impact on ranking for linear walking and turning, respectively (Figure 2).

**Figure 2:**
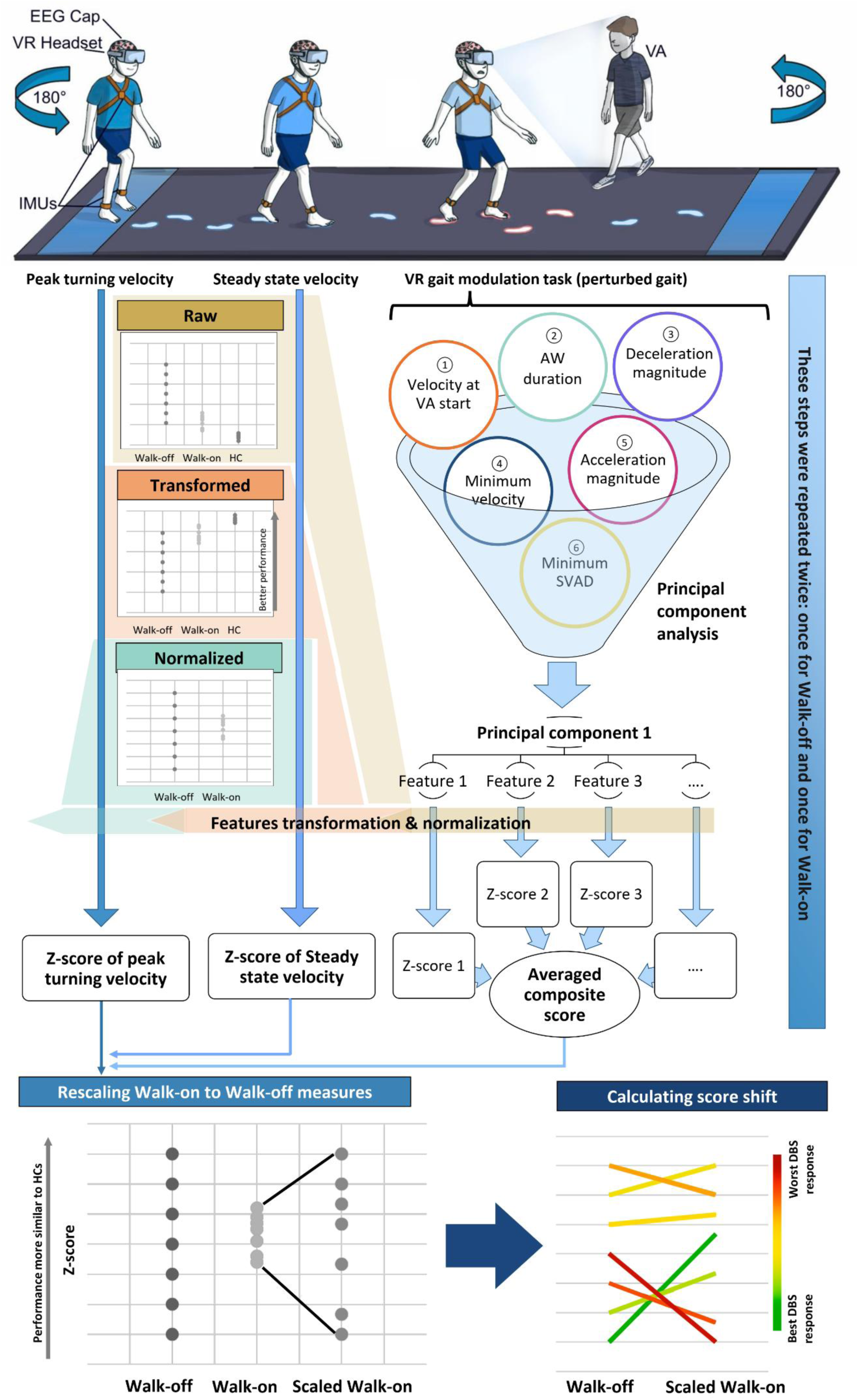
Workflow for deriving and comparing kinematic gait metrics. The schematic illustrates the experimental procedure and analysis pipeline. The upper panel depicts a subject performing the virtual reality (VR) gait adaptation task while equipped with a VR headset, wireless EEG device, and three inertial measurement units (IMUs) placed on the ankles and sternum. Kinematic measures were categorized into two types: (1) metrics unrelated to the perturbation task (i.e., steady-state velocity and peak turning velocity) and (2) metrics capturing responses to the perturbation (i.e., deceleration and acceleration magnitude, minimum and approach velocity, adaptation window [AW] duration, and minimum subject-virtual agent distance [SVAD]). The six features relevant to the perturbation task were subjected to principal component analysis (PCA). Features loading on significant PCs were transformed, normalized (Z-scored), and then averaged to generate a composite gait-adaptation score. Parameters not related to the perturbation (i.e., peak turning velocity and steady-state velocity) were transformed and Z-scored separately. Previous steps were repeated for each experimental condition (Walk-off and Walk-on). To facilitate direct comparison, Walk-on scores were rescaled to the Walk-off score range. Finally, score improvement between Walk-on and Walk-off conditions were calculated to assess DBS-related relative improvement in gait performance.

### 3.4. Brain metabolic imaging acquisition and analysis

A total of 36 FDG-PET scans were acquired in all participants, each undergoing the three conditions (Resting, Walk-off, and Walk-on). Imaging was performed using a Biograph mCT 64 PET/CT scanner (Siemens Medical Solutions). PET data were acquired in 3D mode for 10 min in a single bed position using a 400×400 matrix, with axial resolution of 2 mm full width at half maximum (FWHM) and in-plane resolution of 4.7 mm. Low-dose CT was used for attenuation correction. PET data were iteratively reconstructed in HD mode (24 subsets, three iterations) with Gaussian filtering. Additional scanner details and settings have been reported previously.^47,53,54^

Using statistical parametric mapping (SPM12, Wellcome Centre for Human Neuroimaging, University College London), FDG-PET images were co-registered to the corresponding pre-implantation T1-weighted MPRAGE MRI. T1-weighted images were segmented to obtain deformation fields, which were then applied to the co-registered PET images for spatial normalization to MNI space (MNI152NLin2009bAsym) using non-linear transformation (16 iterations) and trilinear interpolation. The normalized PET images were smoothed with a 10-mm FWHM Gaussian kernel.

Voxel-wise metabolic differences among conditions (Resting, Walk-off, Walk-on) were assessed using a multivariate statistical contrast analysis (F-test), followed by paired t-tests for pairwise comparisons (Resting vs Walk-off and Walk-off vs Walk-on). Statistical maps were thresholded at p<0.001 (uncorrected) with a cluster extent ≥50 voxels. Significant clusters were extracted as binary masks in MNI space for subsequent probabilistic tractography (see Section 3.5).

To examine the relationship between regional metabolic activity and kinematic performance, metabolic measures were extracted from significant SPM clusters using the WFU PickAtlas toolbox. Spherical volumes of interest (VOIs; radius 10 mm) were generated around the peak voxel of each cluster, and contralateral mirrored VOIs were created to assess hemispheric asymmetry. From FDG-PET images, standardized uptake values (SUV) were extracted for each VOI and normalized to the whole-cortex uptake to obtain standardized uptake value ratios (SUVr).

Because all significant clusters were unilateral or showed marked laterality, hemispheric asymmetry was quantified using an asymmetry index, as previously defined,^55,56^ according to the formula:

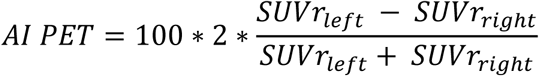

Positive values indicate relatively higher uptake in the left hemisphere, whereas negative values indicate higher uptake in the right hemisphere.

DBS-related changes in metabolic asymmetry between Walk-off and Walk-on conditions were defined as follows:

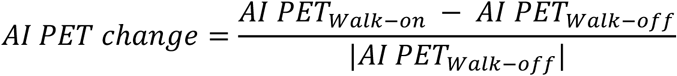

Positive values indicate stimulation-related relative increases in left-hemisphere uptake, whereas negative values reflect greater DBS-related increases in the right hemisphere.

### 3.5. Leads localization and probabilistic tractography analysis

Electrode localization and stimulation site reconstruction were performed using Lead-DBS.^57^ Preoperative T1-weighted MPRAGE MRI images were co-registered with postoperative CT scans, and normalized to MNI space (MNI152NLin2009bAsym) using Advanced Normalization Tools (ANTs) with the DISTAL minimal atlas for standardized anatomical referencing across subjects.^58–60^ Lead reconstruction was then performed using the TRAC/CORE algorithm.^57^ Finally, a 3D visualization was produced using the Lead-Group function.^61^ Accurate localization of active contacts was confirmed for all subjects.

The volume of tissue activated (VTA) was modeled based on the individual stimulation parameters using finite-element method (FEM) electric-field simulations in Lead-DBS. For each subject, the proportion of VTA volume intersecting the STN and its motor subregion was quantified in the MNI space.^59^ The AI of this intersection was calculated:

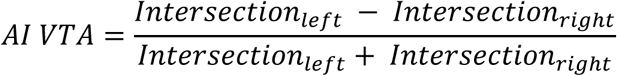

Binarized VTAs were also used to investigate structural connectivity between stimulation sites and PET clusters showing significant metabolic differences between Walk-on and Walk-off conditions. Because DTI was not acquired preoperatively, a normative connectome from 150 unrelated subjects from the Human Connectome Project (HCP) (https://www.humanconnectome.org/) was used for probabilistic tractography.^62^ HCP acquisition and pre-processing have been described previously.^63^

For connectivity analysis, binary masks of bilateral VTAs were used as seeds and significant PET clusters as targets. The masks of the significant clusters were mirrored for the contralateral VTAs to test each VTA connectivity with its ipsilateral targets. After co-registering to diffusion space, a multi-fiber diffusion model was fitted on HCP data using FMRIB’s Diffusion toolbox (FDT) in FMRIB Software Library (FSL, ver. 5.0: http://fsl.fmrib.ox.ac.uk/fsl).^64,65^ This model uses Bayesian techniques to estimate a probability distribution function (PDF) on the principal fiber direction at each voxel, accounting for the possibility of crossing fibers within each voxel. Three fibers were modeled per voxel with a multiplicative factor of one for the prior on the additional modeled fibers and 1000 iterations before sampling.^66^

Using the estimated PDFs and the PROBTRACKX function,^63,67^ the probability of connectivity between the seeds and the targets was determined. From each voxel in the seed mask, the following parameters were used: 5000 streamlines, a 0.2 curvature threshold, and a loop check termination. Target masks were specified as waypoint masks to discard tracts not passing through the target, as termination masks to stop the pathways upon reaching the target, and as classification masks to quantify connectivity values between the seed and the target.^65^ To quantify the strength of connectivity, we used the probability of connectivity, defined as the probability that a particular voxel lies on an existing tract.^68^ For each seed, the probability of connectivity to each target was calculated by dividing the number of streamlines connecting the seed to the target by the total number of streamlines sent out (5000 per seed voxel),^69,70^ according to the following formula, corrected for the seed-to-target distance:

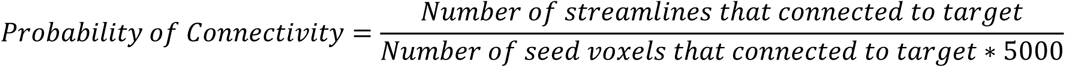

To evaluate the asymmetry of this structural connectivity measure between the two hemispheres, an AI of the tracts was calculated:

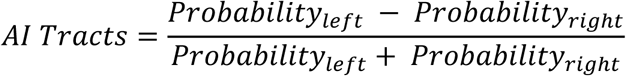

### 3.6. Cortical electrophysiological activity

Three participants (BOS03, PER19, PER20) were excluded because an insufficient number of trials was available for the EEG analysis. For the walking sessions, only trials included in the kinematic analysis were retained for EEG evaluation. In the Resting condition, recordings were segmented into consecutive 30-s epochs and analyzed using the same approach as for the walking sessions.

EEG recordings were pre-processed using a standardized pipeline including: (i) re-referencing to the average of all electrodes; (ii) band-pass filtering in the range of 1–70 Hz using a finite impulse response (FIR) filter with a Hamming window, and a 50 Hz notch filter to suppress power line interference; (iii) Independent Component Analysis (ICA, infomax algorithm) to identify and remove artifacts; (iv) re-referencing to the average to account for residual artifacts introduced by ICA, and (v) Z-score normalization across the entire recording to ensure comparability across sessions and subjects.

The time-frequency representation (TFR) of the cleaned EEG signal was computed for each trial and channel using Morlet wavelet convolution, yielding temporal resolution Δ*t* = 1/2*π s* and frequency resolution Δ*f* = 1 *Hz* across all frequencies. The power spectrum (PS) was then obtained by computing the median of the TFR over time for each trial and channel and then normalized by total power within 1–47 Hz (computed via numerical integration using the trapezoidal rule).

To disentangle oscillatory and aperiodic components, the median PS of all trials and channels was calculated for each condition (Resting, Walk-off, Walk-on), normalized to total power (1–47 Hz), and the aperiodic component was estimated using the FOOOF (Fitting Oscillations & One Over F) algorithm (version 1.1.1). This component was subtracted from the PS of each trial and channel, within each condition. The power within canonical frequency bands (delta: 1-4 Hz, theta: 4-8 Hz, alpha: 8-12 Hz, low beta: 12-21 Hz, high beta: 21-30 Hz, gamma: 30-47 Hz) was then computed as the median over the respective frequency bins. Finally, to obtain a robust summary measure per subject and condition, median PS across trials was computed for each channel.

EEG analyses were performed in Python (version 3.9.19). According to the PET results (see “*4.3 Metabolic signatures of gait adaptation”* section), EEG analysis focused on the F1 and F2 electrodes, corresponding primarily to the left and right SFG, respectively. Task- and stimulation-related power changes were computed as absolute differences between Resting, Walk-off, and Walk-on conditions.

Hemispheric asymmetry of EEG power was quantified as the absolute difference between left and right power spectra (AI EEG) for each frequency band. DBS-related changes in EEG asymmetry were then calculated similarly to the AI PET:

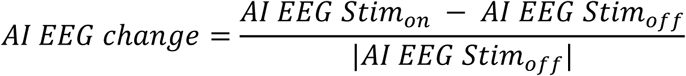

### 3.7. Statistical analysis

DBS-related MDS-UPDRS-III scores improvement was investigated using paired-samples Student T test.

To assess the clinical relevance of the kinematic features, we separately examined the correlations between each of them and the MDS-UPDRS-III scores using Pearson’s correlation. Changes in the individual kinematic features between Walk-off and Walk-on conditions were assessed using the paired Wilcoxon signed-rank test, while differences from the normative range of HC were evaluated using the unpaired Kruskal-Wallis test followed by Bonferroni-corrected post-hoc comparisons.

Pearson’s correlation analysis was also conducted to examine the relationships between two sets of variables: (i) Composite gait-adaptation scores and the Z-scores of SSV and turn peak angular velocity under both Walk-off and Walk-on conditions, and (ii) FDG-PET and EEG measures, the normalized intersection volume between the STN and the VTA, and the probability of connectivity between the VTAs and the SFG.

To identify independent asymmetry-related predictors of DBS-induced changes in the composite gait-adaptation score and brain metabolic activity, we first performed Pearson’s correlation analyses with AI Clinic, AI Stim, AI VTA, and AI Tracks. Asymmetry features not significantly associated with the outcomes were excluded, and the remaining variables were subsequently entered into two separate backward stepwise linear regression models, with DBS-related changes in the composite gait-adaptation score and brain metabolic activity as the respective dependent variables.

Finally, linear regression analysis was also used to isolate the contribution of PET and EEG measures towards the DBS-induced changes in the composite gait-adaptation score.

All statistical analyses were performed using SPSS (version 23.0; IBM/SPSS Inc., Chicago, IL, USA). Significant threshold was set at P=0.05 for all analyses, corrected for multiple comparisons when appropriate.

## 4. Results

### 4.1. Clinical characteristics and STN-DBS response

The mean total MDS-UPDRS-III score in the meds-off/stim-off condition was 44.3 ± 15.4. In this condition, all subjects exhibited axial involvement, with a mean axial MDS-UPDRS-III sub-score of 5.6 ± 3.1. Ten subjects were classified as stage two on the H&Y scale, whereas two (i.e., PER19 and PER22) were classified as stage three. All participants showed a significant response to STN-DBS, with a reduction in the MDS-UPDRS-III total score >10 points, indicating a clinically meaningful improvement, exceeding the threshold for a large clinically important difference.^71^ Axial symptoms also improved during STN-DBS, decreasing to 3.5 ± 2.2 (P < 0.001). In Supplementary tables 1 and 2, we report the demographic, clinical, and STN-DBS programming data for each patient.

### 4.2. Kinematic assessment of gait

The first principal component (PC1) derived from the PCA explained 71.0% and 64.6% of the variability in the Walk-off and Walk-on conditions, respectively. In both conditions, PC1 was primarily loaded by AW duration, average deceleration magnitude, average acceleration magnitude, and velocity at VA onset (Supplementary table 3). As shown in Figure 3, subjects with PD exhibited significant alterations in these four kinematic features relative to HC (Supplementary table 4) in both the Walk-off and Walk-on conditions, with Walk-on values shifting toward those observed in HC. Specifically, AW duration was reduced (mean difference [MD] = –17.5 ± 18.6%, P = 0.010), while deceleration (MD = +92.7 ± 205.5%, P = 0.016), acceleration (MD = +79.5 ± 97.4%, P = 0.010), and velocity at VA onset (MD = +47.5 ± 105.2%, P = 0.006) increased significantly (Supplementary table 5). Features contributing to PC1 as well as the composite gait-adaptation score resulting from them showed statistically significant correlations with the axial and PIGD sub-scores, whereas correlations with the total MDS-UPDRS-III score were weaker, and no significant correlations were observed with the bradykinesia, rigidity, or tremor sub-scores in either the Walk-off or Walk-on conditions (Supplementary table 6). The second principal component accounted only for 23.6% and 30.2% of the variance in Walk-off and Walk-on, respectively, and primarily reflected the interpersonal distance between the subject and the VA (Supplementary table 3).

**Figure 3:**
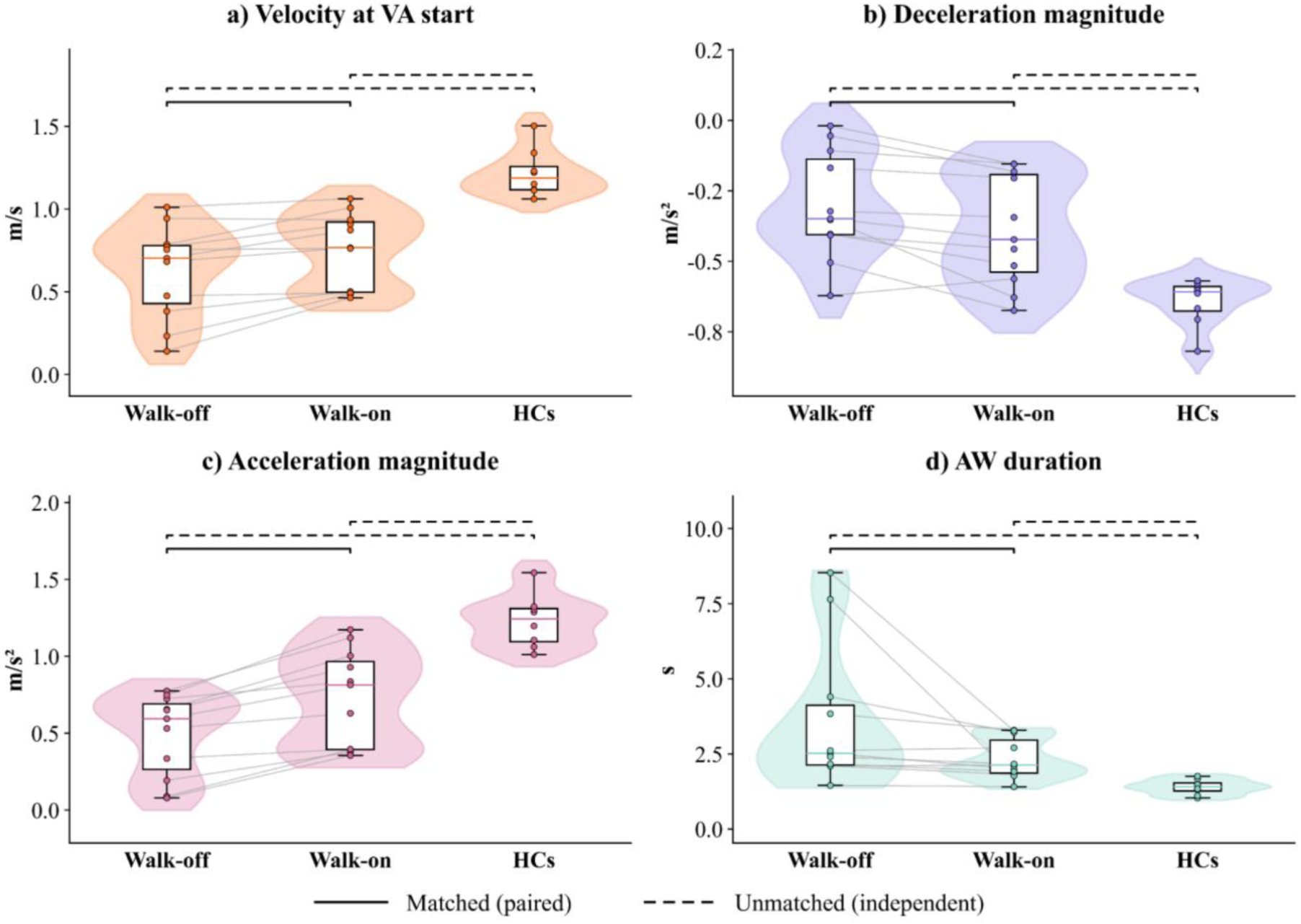
Kinematic variables contributing to the first principal component in subjects with and without stimulation and age-matched healthy controls. The dotted black horizontal lines refer to P < 0.05 in unpaired Kruskal-Wallis statistical test after Bonferroni correction for multiple comparisons; the continuous black horizontal lines refer to P < 0.05 in paired-samples Wilcoxon rank statistical test for the difference from Walk-off to Walk-on after Bonferroni correction for multiple comparisons.

### 4.3. Metabolic signatures of gait adaptation

When comparing Walk-off with Resting condition, we observed increased metabolic activity in the cerebellum (right lobe and vermis), as well as in the sensorimotor cortex (bilateral paracentral lobule and the left precuneus). Conversely, reduced metabolism was observed in the bilateral medial orbitofrontal gyrus (Figure 4 and Supplementary table 7).

**Figure 4:**
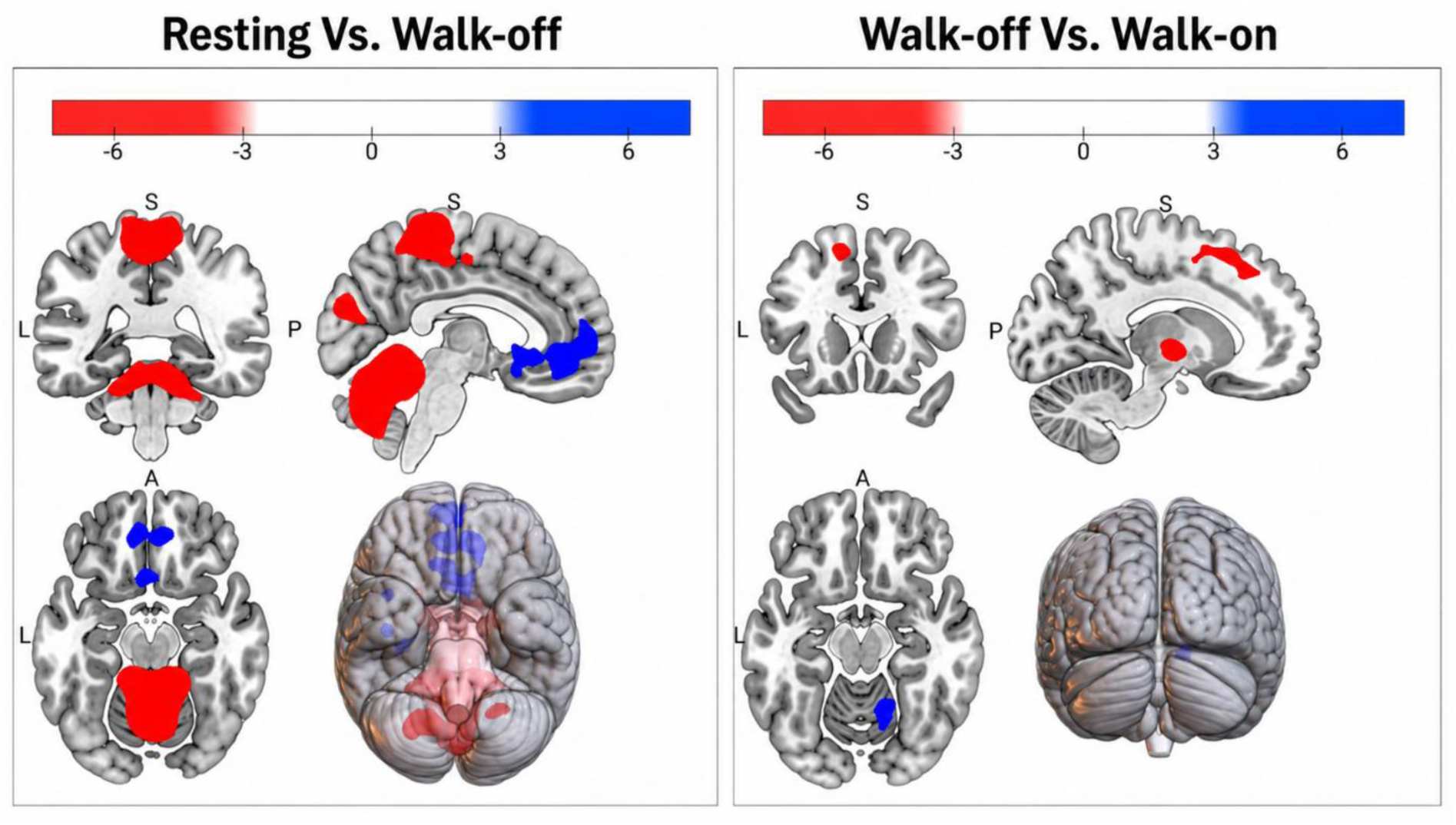
Induced metabolic changes in relation to the performance of the task and the activation of DBS. The left panel shows the contrast findings between the FDG-PET scan performed during resting and obstacle avoidance in stim-off (Resting vs. Walk-off); Right panel shows the contrast findings between the scans performed during obstacle avoidance in stim-off and stim-on (Walk-off vs. Walk-on). Red: higher uptake; Blue: lower uptake; Further details regarding the statistical analysis behind the images can be found in Supplementary Table 7.

Activation of DBS during walking (Walk-on vs. Walk-off) was associated with increased metabolism in the left SFG and in the left thalamus, alongside decreased activity in the right cerebellum (Figure 4 and Supplementary table 7).

No significant correlations were observed between SUVr values and the corresponding composite gait-adaptation scores in both the Walk-off and Walk-on conditions.

The AI PET in the SFG instead showed significant negative correlation with the composite score in both the Walk-off (r = –0.685, P = 0.020) and Walk-on (r = –0.689, P = 0.019) conditions. This finding indicates that a relatively higher SUVr in the right SFG compared to the left is associated with better gait adaptation performance, independent of the DBS status. The AI PET change also showed a negative correlation with DBS-induced improvement in the composite score (r = –0.683, P = 0.020). Specifically, subjects whose DBS settings induced relatively greater metabolic activity in the right SFG compared to the left SFG demonstrated greater improvements in gait adaptation performance (Figure 5).

**Figure 5:**
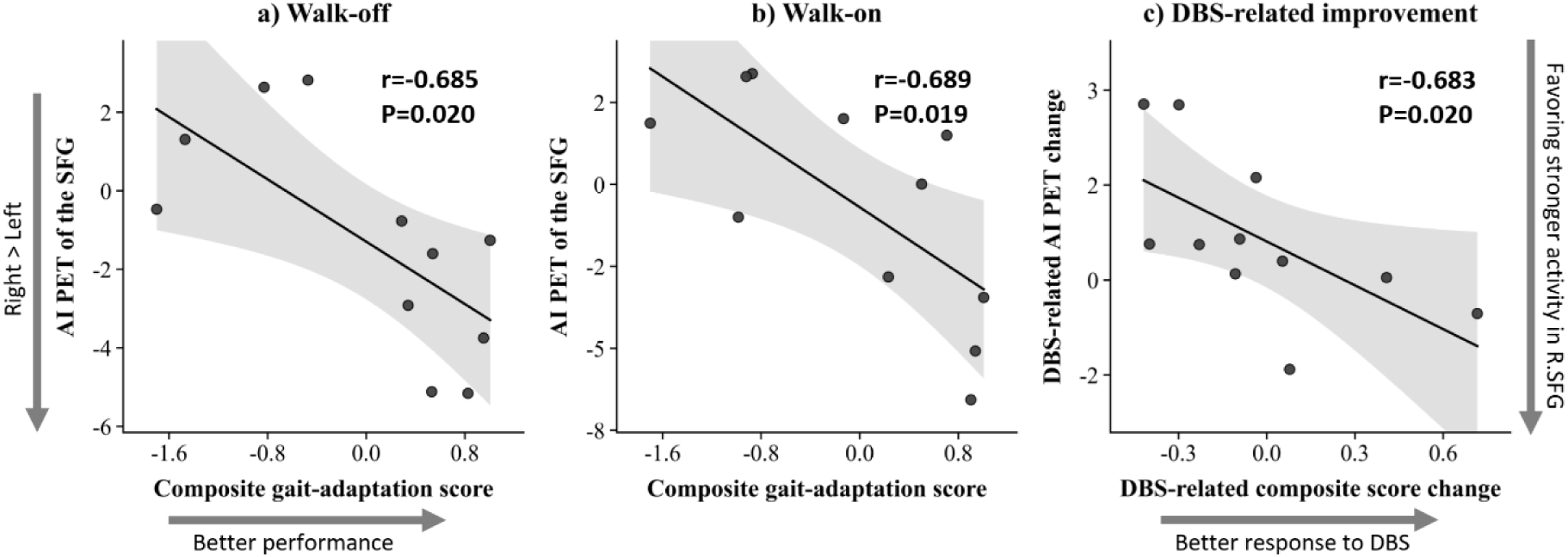
Correlation between the asymmetry in the metabolic activity in the SFG and the composite gait-adaptation score. AI PET: asymmetry index of the SUVrs between VOIs of the left and right SFG, where its positive and negative values demonstrate greater relative SUVR in the left and right hemisphere, respectively; DBS-related AI PET change values are positive when DBS favors higher SUVr in the left SFG, and negative when it favors higher SUVr in the right SFG; r: Pearson correlation coefficient.

Backward stepwise linear regression, with DBS-induced change in the composite score as the dependent variable and AI Clinic, AI Stim, and AI PET change as predictors, identified AI PET change in the SFG as the sole independent predictor (Odds Ratio [OR] = -0.683, 95% CI [-0.327, -0.035], R² = 0.408, P = 0.020). In a separate backward stepwise linear regression analysis with AI PET change in the SFG as the dependent variable and AI Clinic and AI Stim as predictors, AI Stim emerged as the only independent predictor (OR = -0.725, 95% CI [-11.354, -1.884], R² = 0.474, P = 0.012; Supplementary table 8), excluding a potential confounding effect on these results by the distribution of akinetic-rigid symptoms between the two hemibodies. The effect of other possible confounding factors such as differences in lead localization and STN-DBS programming across hemispheres was also excluded. Specifically, the MDS-UPDRS-III sub-scores for bradykinesia, rigidity, and tremor, assessed for each hemibody under both stimulation conditions and single-side stimulation intensity were not correlated with DBS-related changes in the composite score (data not shown). We did not observe differences in lead placement or active contact locations among patients with low, moderate, and high levels of improvement (Supplementary figure 1). Tracts connecting the VTAs to the SFG clusters showed substantial overlap, with no significant differences across the three groups (Supplementary figure 2). No significant correlations were found between DBS-induced gait improvement and AI VTA (Supplementary figure 3) or AI Tracks (Supplementary figure 4; Supplementary table 9).

To assess the specificity of the AI PET SFG to gait adaptation performance, we examined the same correlations using two control variables: SSV and peak turning velocity (Figure 2). Both control variables significantly increased in the Walk-on condition compared to Walk-off (Supplementary Table 10). While their values significantly correlated with the AI PET in both stimulation conditions, their DBS-induced changes did not correlate with the DBS-related change in the AI PET (Supplementary Figure 5).

### 4.4. Cortical electrophysiological activity relationship with FDG uptake and gait improvement

Power spectral density analysis revealed no significant differences across conditions in any frequency band and no correlations with SUVr (data not shown).

In the theta-band, DBS-related AI EEG change in the SFG showed a significant negative correlation with changes in the composite gait-adaptation score (r = -0.753, P = 0.031), mirroring the relationship observed for AI PET change (Figure 6). The two metrics were however not correlated (r = 0.284, P = 0.495).

**Figure 6:**
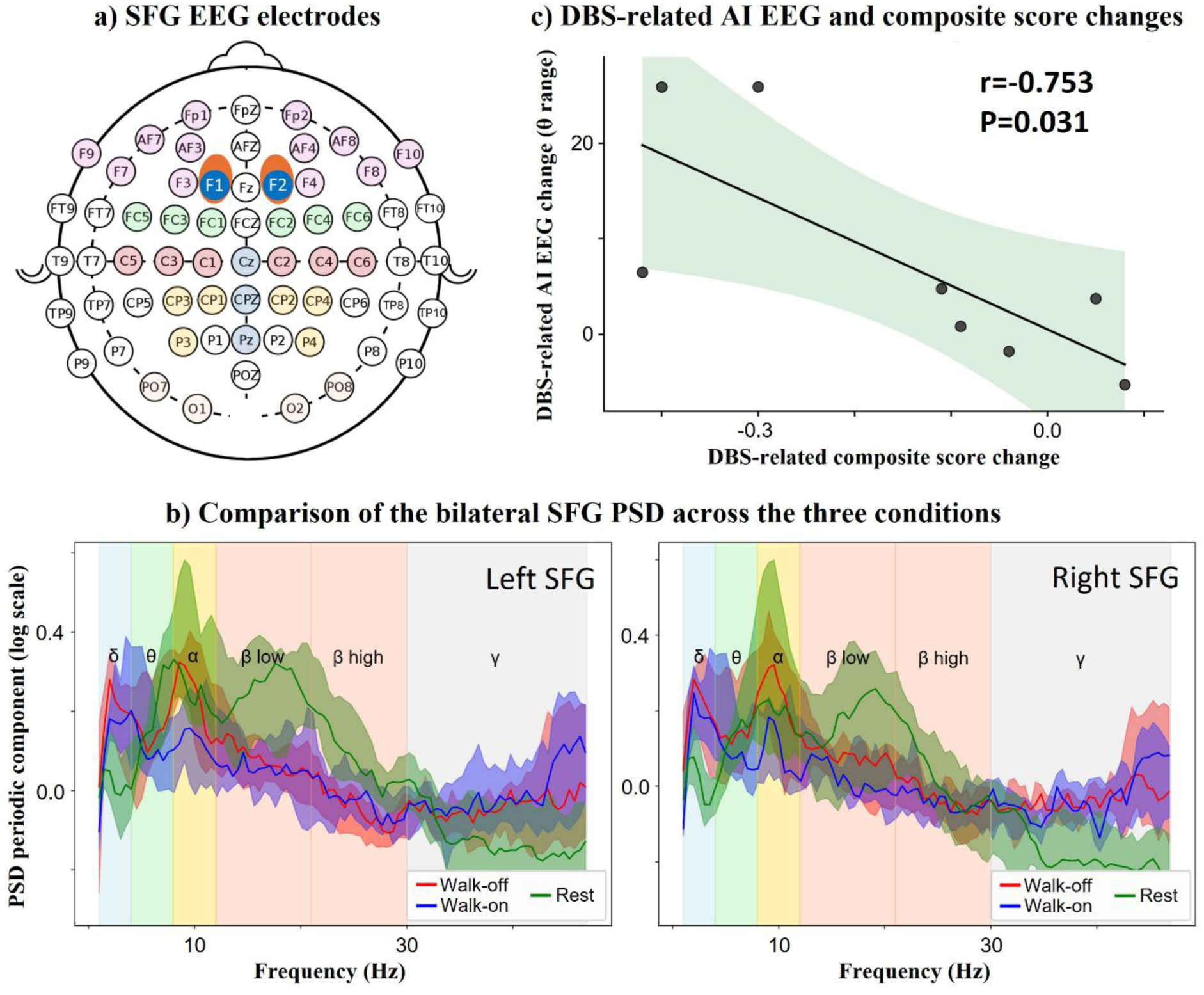
DBS-induced changes in the asymmetry of EEG activity in the SFG. (a) Electrode layout of the EEG cap used in the study (the orange areas refer to the SFG region); (b) Bilateral PSD comparisons across the three experimental conditions; no significant differences were found in any band; (c) Negative correlation between DBS-induced changes in AI EEG in the SFG (theta band) and changes in the composite gait-adaptation score. AI EEG: asymmetry index of EEG band power between electrodes over the left and right SFG; DBS-related changes in AI EEG are positive when DBS favors greater power in the left SFG and negative when DBS favors greater power in the right SFG; r: Pearson correlation coefficient.

A linear regression model incorporating AI EEG change and AI PET change significantly predicted improvement in the composite score (F = 9.815, R² = 0.797, P = 0.019). In this model, both predictors contributed independently: AI EEG (B = -0.010 [-0.019, -0.001], P = 0.034) and AI PET change (B = -0.072 [-0.151, 0.006], P = 0.063).

## 5. Discussion

Our findings identify a lateralized thalamo–frontal network with prominent involvement of the SFG that masters gait adaptation in PD. By integrating kinematic, metabolic, and electrophysiological data acquired during an ecologically valid walking task, we show that STN-DBS improves adaptive locomotion by functioning as an active control mechanism that dynamically reallocates network activity across hemispheres.

The kinematic composite score developed in this study captures the fundamental dynamics of obstacle avoidance, including approach velocity, gait adaptation duration, and acceleration-deceleration capacity. These parameters were markedly altered in patients with PD, reflecting reduced locomotor flexibility and impaired scaling of motor output. Such deficits are consistent with previous studies showing impaired anticipatory postural adjustments and reduced braking capacity during gait adaptation in PD.^20,72–76^ Importantly, these kinematic features correlated specifically with axial and PIGD symptoms, suggesting that they capture locomotor deficits that are partially reflected in conventional clinical scales.^77,78^ Activation of STN-DBS significantly improved these measures toward the normative range observed in healthy participants. This finding indicates that STN-DBS can partially restore adaptive locomotor control in PD.

The gait-adaptation task engaged a distributed locomotor network including the cerebellum and sensorimotor cortex. Increased cerebellar metabolism during walking likely reflects compensatory recruitment of cerebello-cortical circuits in response to basal ganglia dysfunction, a pattern consistently observed in imaging studies of parkinsonian gait.^47,79,80^ Notably, cerebellar activity decreased during STN-DBS in parallel with improved gait performance. This reduction suggests that effective STN-DBS may restore more efficient basal ganglia-thalamo-cortical processing, thereby reducing the need for compensatory cerebellar involvement.^80,81^

The most behaviorally relevant metabolic effects of STN-DBS on adaptive gait involved the SFG and thalamus. The thalamus is known to function as an integrative relay that conveys motor, cognitive, and limbic information essential for the coordination and context-appropriate control of gait.^1,82^ The SFG is a key node within prefrontal-basal ganglia circuits that supports executive aspects of motor control, including action selection and the monitoring of environmental constraints.^83–86^ Notably, gait-adaptation performance correlated with hemispheric asymmetry of SFG metabolism rather than with absolute activity levels. Greater right-hemispheric dominance of SFG activity was associated with better gait performance. This interpretation aligns with experimental and clinical evidence implicating right frontal cortical regions in action inhibition, behavioral monitoring, and motor control under uncertainty.^29,87,88^ Disruption of this network has been associated with impulsive motor behavior and impaired adaptive control.^29,89,90^

Electrophysiological recordings provided additional evidence that STN-DBS modulates lateralized frontal network dynamics during gait adaptation. Specifically, DBS-induced changes in theta-band asymmetry over frontal electrodes were significantly associated with improvements in gait adaptation performance. Theta-band activity within frontal cortical regions has been consistently linked to cognitive control, conflict monitoring, and adaptive motor behavior, particularly in contexts requiring rapid updating of motor plans.^6,91^ In the present study, greater rightward shifts in frontal theta activity paralleled improvements in gait adaptation, supporting the interpretation that adaptive locomotion depends on the integrity of lateralized frontal control processes. Importantly, these electrophysiological changes mirrored the metabolic asymmetry observed in the SFG, despite the absence of a direct correlation between EEG and PET measures. This apparent dissociation likely reflects the different temporal scales captured by the two modalities, with EEG indexing fast neural dynamics and PET reflecting slower metabolic processes. Nevertheless, the convergence of both measures on a lateralized frontal mechanism strengthens the interpretation that STN-DBS reshapes functional network balance rather than acting locally at the level of the STN. Furthermore, it offers a practical and accessible clinical tool for monitoring DBS-related frontal asymmetry, without the need for invasive or expensive neuroimaging.

Taken together, these findings support a model in which STN-DBS acts as a lateralized regulator of distributed network activity. Rather than uniformly enhancing or suppressing cortical activity, STN-DBS appears to dynamically reallocate neural activity within bilateral frontal-basal ganglia circuits, shifting the relative contribution of each hemisphere to gait adaptation. Indeed, the asymmetry of STN-DBS amplitude was the single independent predictor of the lateralization in FDG-PET, which dictated the level of gait adaptation improvement.

A plausible mechanism underlying this effect involves modulation of cortico-subthalamic projections within the hyperdirect pathway, which provides a rapid excitatory route for top-down control of motor output.^92–94^ This pathway is critically involved in action inhibition and rapid adjustment of behavior, functions that are essential for adaptive gait.^92,95^ Within this framework, STN-DBS may alter locomotor behavior by modulating the balance of inhibitory control exerted by bilateral frontal regions over subcortical motor circuits. In particular, our findings suggest that preserving right-hemispheric frontal control by delivering less relative stimulation to the right STN is associated with greater effectiveness of STN-DBS on gait adaptation in PD (Figure 7). Collectively, these data support a model in which STN-DBS does not simply modulate motor output but dynamically reconfigures the balance of activity within lateralized frontal control networks.^96^ In this model, optimal gait adaptation emerges from a coordinated interaction between thalamocortical drive and right-dominant frontal inhibitory control, allowing flexible adjustment of locomotor behavior in response to environmental demands.

**Figure 7:**
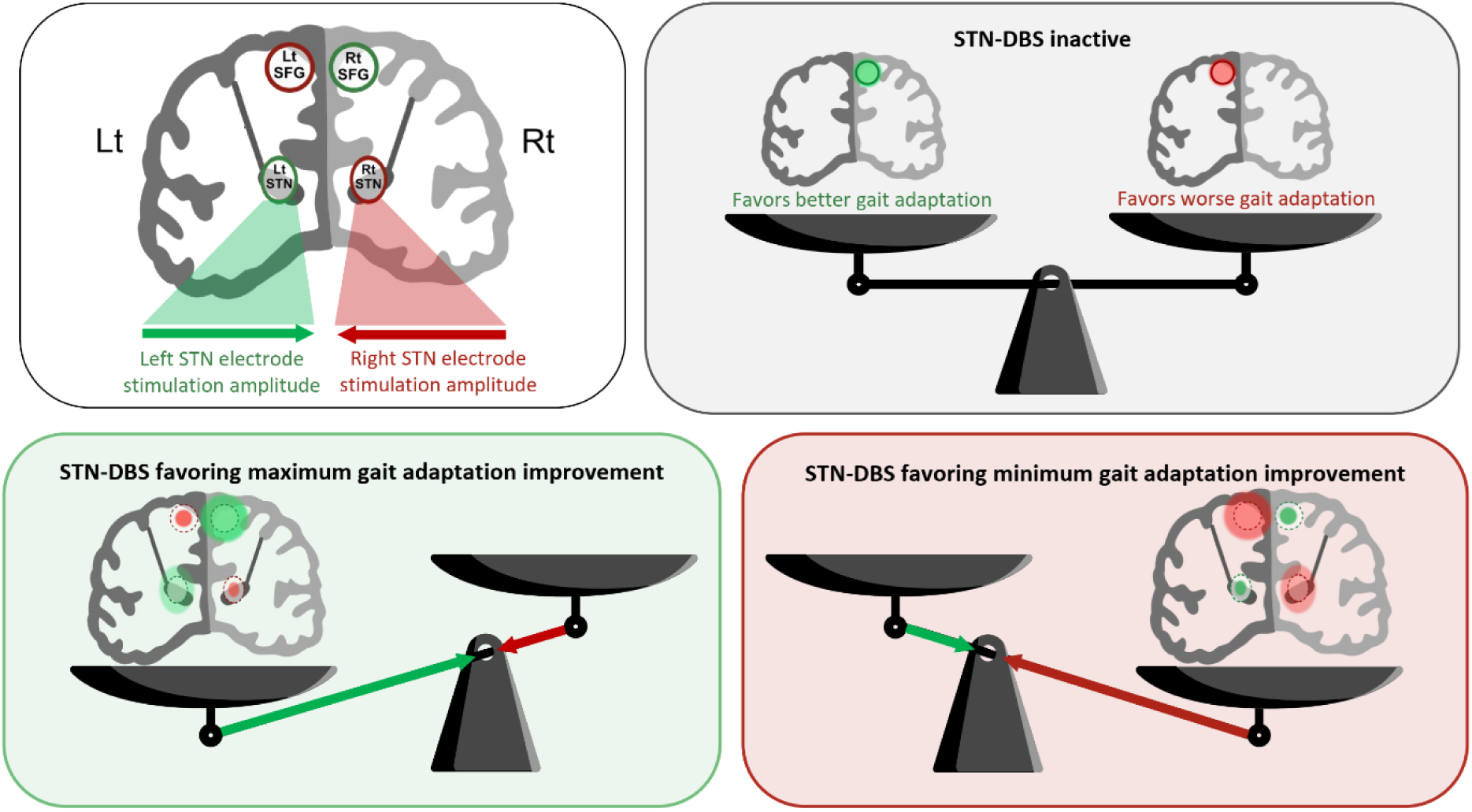
Conceptual model linking gait adaptation performance to its neural underpinnings, highlighting DBS as a lateralized regulator of brain activity that influences gait adaptation improvement. The upper left panel provides a schematic legend of activity within the left and right superior frontal gyri (Lt SFG and Rt SFG, respectively), with arrows additionally indicating the amplitude of deep brain stimulation (DBS) delivered to the left and right subthalamic nuclei (Lt STN and Rt STN, respectively). The upper right panel illustrates effect of asymmetric SFG activity on gait adaptation performance under inactive STN-DBS. The lower panels depict how asymmetry in STN-DBS stimulation amplitude influences asymmetric SFG activation, thereby promoting either maximal (lower left panel) or minimal (lower right panel) DBS-related improvement in gait adaptation.

### 5.1. Clinical implication and limitations

These findings have several potential clinical implications. First, they suggest that gait disturbances in PD may arise not only from impaired motor output but also from disruption of frontal control systems regulating adaptive locomotion. Second, they indicate that the efficacy of STN-DBS for gait disturbances may depend on its ability to rebalance activity within lateralized frontal-basal ganglia networks. Third, the identification of SFG asymmetry as a predictor of gait improvement raises the possibility that hemispheric balance within frontal networks could serve to optimize image-guided DBS programming strategies.

Several limitations should be acknowledged. The relatively small sample size reflects the methodological complexity of combining immersive locomotor paradigms with multimodal imaging; nevertheless, the convergence of independent modalities supports the robustness of the findings. The exclusive inclusion of right-handed participants limits the generalizability of hemispheric effects, which may relate to dominant/non-dominant organization rather than strictly anatomical laterality. However, the improvement in gait adaptation was not a function of higher stimulation amplitude on one hemisphere, nor on the underlying baseline symptom asymmetry, or subject’s baseline asymmetry in motor impairment. Additionally, five patients experienced freezing of gait episodes in the Walk-off condition. Separate analysis of these episodes was not feasible due to the limited sample size and the inability of FDG-PET to temporally resolve episodic gait disturbances such as freezing of gait during the FDG uptake period. Lastly, the use of normative structural connectivity data may not fully capture individual variability in white matter pathways.

### 5.2. Conclusion

This study provides convergent behavioral, metabolic, and electrophysiological evidence that adaptive locomotion in PD depends on a lateralized frontal-basal ganglia network centered on the SFG. STN-DBS may improve gait adaptation by modulating lateralized cortical engagement, shifting frontal cortical dynamics toward a configuration that determines the efficacy of top-down control of locomotion. These findings support a network-level model of DBS action and highlight hemispheric balance within frontal control circuits as a potential determinant of therapeutic efficacy for gait disturbances in PD.

## 6. Data availability statement

The data supporting this manuscript is available upon request to the corresponding author.

## 7. Acknowledgement

The authors wish to express their gratitude to Ghadir Abbas, Aya Alhabbal, Aya Alsharif, Joachim Brumberg, Philip Capetian, Kheder Kheder, Manuel Kunzig, Maarouf Gorra Al Nafouri, Martin Reich, Simone Seifert for their valuable assistance in data acquisition. We are also thankful to Suzana Sultan for her support in data visualization. The authors used OpenAI tools solely for language and stylistic editing of the manuscript; all content was critically reviewed and approved by the authors

## 10. Supplementary material

Supplementary material is available at *Brain* online.

## 11. Authors’ contributions

Conceptualization, N.G.P., I.U.I., C.P.; methodology, N.G.P., G.M., I.U.I., C.P.; formal analysis, I.H.; R.H., S.F., G.M., L.R., C.P., investigation, I.H., N.G.P., I.U.I., C.P.; resources, I.U.I., C.P.; data curation, I.H., P.C.; writing – original draft preparation, I.H., N.G.P., S.F., G.M., L.R., I.U.I., C.P.; writing – review and editing, A.B., G.P., J.V.; visualization, I.H., C.P.; supervision, I.U.I.; project administration, I.U.I.; funding acquisition, I.U.I., C.P.. All authors have read and agreed to the published version of the manuscript.

## References

1. Takakusaki K. Functional Neuroanatomy for Posture and Gait Control. Journal of Movement Disorders. 2017;10(1):1–17. doi:10.14802/jmd.16062

2. Smulders K, Dale ML, Carlson-Kuhta P, Nutt JG, Horak FB. Pharmacological treatment in Parkinson’s disease: Effects on gait. Parkinsonism & Related Disorders. 2016;31:3–13. doi:10.1016/j.parkreldis.2016.07.006

3. Pozzi NG, Palmisano C, Reich MM, et al. Troubleshooting Gait Disturbances in Parkinson’s Disease With Deep Brain Stimulation. Frontiers in Human Neuroscience. 2022;Volume 16-2022. doi:10.3389/fnhum.2022.806513

4. Roemmich RT, Hack N, Akbar U, Hass CJ. Effects of dopaminergic therapy on locomotor adaptation and adaptive learning in persons with Parkinson’s disease. Behavioural Brain Research. 2014;268:31–39. doi:10.1016/j.bbr.2014.03.041

5. Pieruccini-Faria F, Vitório R, Almeida QJ, et al. Evaluating the Acute Contributions of Dopaminergic Replacement to Gait With Obstacles in Parkinson’s Disease. https://doi.org/1101080/002228952013810139. 2013;45(5):369–380. doi:10.1080/00222895.2013.810139

6. Pozzi NG, Canessa A, Palmisano C, et al. Freezing of gait in Parkinson’s disease reflects a sudden derangement of locomotor network dynamics. Brain. 2019;142(7):2037–2050. doi:10.1093/brain/awz141

7. Pottorf TS, Nocera JR, Eicholtz SP, Kesar TM. Locomotor Adaptation Deficits in Older Individuals With Cognitive Impairments: A Pilot Study. Frontiers in Neurology. 2022;Volume 13-2022. https://www.frontiersin.org/journals/neurology/articles/10.3389/fneur.2022.800338

8. Mirelman A, Bonato P, Camicioli R, et al. Gait impairments in Parkinson’s disease. The Lancet Neurology. 2019;18(7):697–708. doi:10.1016/S1474-4422(19)30044-4

9. Grimbergen YAM, Schrag A, Mazibrada G, Borm GF, Bloem BR. Impact of Falls and Fear of Falling on Health-Related Quality of Life in Patients with Parkinson’s Disease. Journal of Parkinson’s Disease. 2013;3(3):409–413. doi:10.3233/JPD-120113

10. Müller MLTM, Marusic, Uros, van Emde Boas, Miriam, Weiss, Daniel, and Bohnen NI. Treatment options for postural instability and gait difficulties in Parkinson’s disease. Expert Review of Neurotherapeutics. 2019;19(12):1229–1251. doi:10.1080/14737175.2019.1656067

11. Hariz M, Blomstedt P. Deep brain stimulation for Parkinson’s disease. Journal of Internal Medicine. 2022;292(5):764–778. doi:10.1111/joim.13541

12. Pötter-Nerger M, Volkmann J. Deep brain stimulation for gait and postural symptoms in Parkinson’s disease. 2013;28(11):1609–1615.

13. van Nuenen BFL, Esselink RAJ, Munneke M, Speelman JD, van Laar T, Bloem BR. Postoperative gait deterioration after bilateral subthalamic nucleus stimulation in Parkinson’s disease. Movement Disorders. 2008;23(16):2404–2406. 10.1002/mds.21986

14. Lohnes CA, Earhart GM. Effect of subthalamic deep brain stimulation on turning kinematics and related saccadic eye movements in Parkinson disease. Experimental Neurology. 2012;236(2):389–394. doi:10.1016/j.expneurol.2012.05.001

15. Hong M, Earhart GM. Effects of Medication on Turning Deficits in Individuals with Parkinson’s Disease. Journal of Neurologic Physical Therapy. 2010;34(1). https://journals.lww.com/jnpt/fulltext/2010/03000/effects_of_medication_on_turning_defi cits_in.3.aspx

16. Curtze C, Nutt JG, Carlson-Kuhta P, Mancini M, Horak FB. Levodopa is a Double-Edged Sword for Balance and Gait in People with Parkinson’s Disease. Movement disorders : official journal of the Movement Disorder Society. 2015;30(10):1361. doi:10.1002/MDS.26269

17. Pillai L, Landes RD, Virmani T. Levodopa influence on turning dynamics in people with Parkinson’s disease. Scientific Reports. 2025;15(1):14141. doi:10.1038/s41598-025-97511-4

18. Wai Y, Wang J, Weng Y, et al. Cortical involvement in a gait-related imagery task: comparison between Parkinson’s disease and normal aging. Parkinsonism & related disorders. 2012;18(5):537–542. doi:10.1016/J.PARKRELDIS.2012.02.004

19. Ohara M, Hirata K, Hallett M, et al. Long-term levodopa ameliorates sequence effect in simple, but not complex walking in early Parkinson’s disease patients. Parkinsonism & Related Disorders. 2023;108:105322. doi:10.1016/j.parkreldis.2023.105322

20. Orcioli-Silva D, Barbieri FA, Simieli L, et al. Walking behavior over multiple obstacles in people with Parkinson’s disease. Gait and Posture. 2017;58:510–515. doi:10.1016/j.gaitpost.2017.09.021

21. Piet A, Geritz J, Garcia P, et al. Predicting executive functioning from walking features in Parkinson’s disease using machine learning. Scientific Reports. 2024;14(1):29522. doi:10.1038/s41598-024-80144-4

22. Pourahmad R, Saleki K, Esmaili M, et al. Deep brain stimulation (DBS) as a therapeutic approach in gait disorders: What does it bring to the table? IBRO Neuroscience Reports. 2023;14:507–513. doi:10.1016/j.ibneur.2023.05.008

23. Takakusaki K, Takahashi M, Noguchi T, Chiba R. Neurophysiological mechanisms of gait disturbance in advanced Parkinson’s disease patients. Neurology and Clinical Neuroscience. 2023;11(4):201–217. doi:10.1111/ncn3.12683

24. Pelicioni PHS, Lord SR, Okubo Y, Menant JC. Cortical activation during gait adaptability in people with Parkinson’s disease. Gait & Posture. 2022;91:247–253. doi:10.1016/j.gaitpost.2021.10.038

25. Maidan I, Nieuwhof F, Bernad-Elazari H, et al. The Role of the Frontal Lobe in Complex Walking Among Patients With Parkinson’s Disease and Healthy Older Adults: An fNIRS Study. Neurorehabilitation and neural repair. 2016;30(10):963–971. doi:10.1177/1545968316650426

26. Steidel K, Ruppert MC, Palaghia I, et al. Dopaminergic pathways and resting-state functional connectivity in Parkinson’s disease with freezing of gait. NeuroImage: Clinical. 2021;32:102899. doi:10.1016/j.nicl.2021.102899

27. Mosley PE, Robinson K, Coyne T, et al. Subthalamic deep brain stimulation identifies frontal networks supporting initiation, inhibition and strategy use in Parkinson’s disease. NeuroImage. 2020;223:117352. doi:10.1016/j.neuroimage.2020.117352

28. Boisgueheneuc FD, Levy R, Volle E, et al. Functions of the left superior frontal gyrus in humans: a lesion study. Brain. 2006;129(12):3315–3328. doi:10.1093/brain/awl244

29. Hu S, Ide JS, Zhang S, Li C shan R. The Right Superior Frontal Gyrus and Individual Variation in Proactive Control of Impulsive Response. J Neurosci. 2016;36(50):12688. doi:10.1523/JNEUROSCI.1175-16.2016

30. Jin C, Yang L, Qi S, et al. Structural Brain Network Abnormalities in Parkinson’s Disease With Freezing of Gait. Frontiers in Aging Neuroscience. 2022;Volume 14-2022. https://www.frontiersin.org/journals/aging-neuroscience/articles/10.3389/fnagi.2022.944925

31. Bharti K, Suppa A, Tommasin S, et al. Neuroimaging advances in Parkinson’s disease with freezing of gait: A systematic review. NeuroImage: Clinical. 2019;24:102059. doi:10.1016/j.nicl.2019.102059

32. Mi TM, Mei SS, Liang PP, et al. Altered resting-state brain activity in Parkinson’s disease patients with freezing of gait. Scientific Reports. 2017;7(1):16711. doi:10.1038/s41598-017-16922-0

33. Bastian AJ, Kelly VE, Revilla FJ, Perlmutter JS, Mink JW. Different effects of unilateral versus bilateral subthalamic nucleus stimulation on walking and reaching in Parkinson’s disease. Movement Disorders. 2003;18(9):1000–1007. doi:10.1002/mds.10493

34. Roper JA, Kang N, Ben J, Cauraugh JH, Okun MS, Hass CJ. Deep brain stimulation improves gait velocity in Parkinson’s disease: a systematic review and meta-analysis. Journal of Neurology. 2016;263(6):1195–1203. doi:10.1007/s00415-016-8129-9

35. Wang J, Shang R, He L, et al. Prediction of Deep Brain Stimulation Outcome in Parkinson’s Disease With Connectome Based on Hemispheric Asymmetry. Frontiers in Neuroscience. 2021;Volume 15-2021. doi:10.3389/fnins.2021.620750

36. Schott FP, Gulberti A, Pinnschmidt HO, et al. Subthalamic Deep Brain Stimulation Lead Asymmetry Impacts the Parkinsonian Gait Disorder. Frontiers in Human Neuroscience. 2022;Volume 16-2022. doi:10.3389/fnhum.2022.788200

37. Lin Z, Zhang C, Li D, Sun B. Lateralized effects of deep brain stimulation in Parkinson’s disease: evidence and controversies. npj Parkinson’s Disease. 2021;7(1):64. doi:10.1038/s41531-021-00209-3

38. Hughes AJ, Daniel SE, Kilford L, Lees AJ. Accuracy of clinical diagnosis of idiopathic Parkinson’s disease: a clinico-pathological study of 100 cases. Journal of Neurology, Neurosurgery & Psychiatry. 1992;55(3):181–184. doi:10.1136/jnnp.55.3.181

39. Arlotti M, Marceglia S, Foffani G, et al. Eight-hours adaptive deep brain stimulation in patients with Parkinson disease. Neurology. 2018;90(11):e971–e976. doi:10.1212/WNL.0000000000005121

40. Steigerwald F, Pötter M, Herzog J, et al. Neuronal Activity of the Human Subthalamic Nucleus in the Parkinsonian and Nonparkinsonian State. Journal of Neurophysiology. 2008;100(5):2515–2524. doi:10.1152/jn.90574.2008

41. Li X, Xing Y, Martin-Bastida A, Piccini P, Auer DP. Patterns of grey matter loss associated with motor subscores in early Parkinson’s disease. NeuroImage: Clinical. 2018;17:498–504. doi:10.1016/j.nicl.2017.11.009

42. Kelly VE, Johnson CO, McGough EL, et al. Association of cognitive domains with postural instability/gait disturbance in Parkinson’s disease. Parkinsonism & Related Disorders. 2015;21(7):692–697. doi:10.1016/j.parkreldis.2015.04.002

43. Stebbins GT, Goetz CG, Burn DJ, Jankovic J, Khoo TK, Tilley BC. How to identify tremor dominant and postural instability/gait difficulty groups with the movement disorder society unified Parkinson’s disease rating scale: Comparison with the unified Parkinson’s disease rating scale. Movement Disorders. 2013;28(5):668–670. doi:10.1002/mds.25383

44. Seuthe J, Hermanns H, Hulzinga F, et al. Gait asymmetry and symptom laterality in Parkinson’s disease: two of a kind? Journal of Neurology. 2024;271(7):4373–4382. doi:10.1007/s00415-024-12379-0

45. Tomlinson CL, Stowe R, Patel S, Rick C, Gray R, Clarke CE. Systematic review of levodopa dose equivalency reporting in Parkinson’s disease. Movement Disorders. 2010;25(15):2649–2653. doi:10.1002/mds.23429

46. World Medical Association. World Medical Association Declaration of Helsinki: Ethical Principles for Medical Research Involving Human Subjects. JAMA. 2013;310(20):2191–2194. doi:10.1001/jama.2013.281053

47. Pellegrini F, Pozzi NG, Palmisano C, et al. Cortical networks of parkinsonian gait: a metabolic and functional connectivity study. Annals of Clinical and Translational Neurology. 2024;11(10):2597–2608. doi:10.1002/acn3.52173

48. Palmisano C, Kullmann P, Hanafi I, et al. A Fully-Immersive Virtual Reality Setup to Study Gait Modulation. Frontiers in Human Neuroscience. 2022;16:80. doi:10.3389/FNHUM.2022.783452/XML/NLM

49. Parrington L, King LA, Weightman MM, et al. Between-site equivalence of turning speed assessments using inertial measurement units. Gait & Posture. 2021;90:245–251. doi:10.1016/j.gaitpost.2021.09.164

50. El-Gohary M, Pearson S, McNames J, et al. Continuous Monitoring of Turning in Patients with Movement Disability. Sensors. 2014;14(1):356–369. doi:10.3390/s140100356

51. Salarian A, Horak FB, Zampieri C, Carlson-Kuhta P, Nutt JG, Aminian K. iTUG, a Sensitive and Reliable Measure of Mobility. IEEE Transactions on Neural Systems and Rehabilitation Engineering. 2010;18(3):303–310. doi:10.1109/TNSRE.2010.2047606

52. Morris R, Martini DN, Smulders K, et al. Cognitive associations with comprehensive gait and static balance measures in Parkinson’s disease. Parkinsonism & Related Disorders. 2019;69:104–110. doi:10.1016/j.parkreldis.2019.06.014

53. Hinton GE, Salakhutdinov RR. Reducing the Dimensionality of Data with Neural Networks. Science. 2006;313(5786):504–507. doi:10.1126/science.1127647

54. Reich MM, Brumberg J, Pozzi NG, et al. Progressive gait ataxia following deep brain stimulation for essential tremor: adverse effect or lack of efficacy? Brain. 2016;139(11):2948–2956. doi:10.1093/brain/aww223

55. Hong Y, Fu C, Xing Y, et al. Delayed 18F-FDG PET imaging provides better metabolic asymmetry in potential epileptogenic zone in temporal lobe epilepsy. Frontiers in Medicine. 2023;Volume 10-2023. https://www.frontiersin.org/journals/medicine/articles/10.3389/fmed.2023.1180541

56. Jeong Y, Cho SS, Park JM, et al. ^18^F-FDG PET Findings in Frontotemporal Dementia: An SPM Analysis of 29 Patients. J Nucl Med. 2005;46(2):233.

57. Horn A, Kühn AA. Lead-DBS: A toolbox for deep brain stimulation electrode localizations and visualizations. NeuroImage. 2015;107:127–135. 10.1016/j.neuroimage.2014.12.002

58. Avants BB, Tustison NJ, Song G, Cook PA, Klein A, Gee JC. A reproducible evaluation of ANTs similarity metric performance in brain image registration. NeuroImage. 2011;54(3):2033–2044. 10.1016/j.neuroimage.2010.09.025

59. Ewert S, Plettig P, Li N, et al. Toward defining deep brain stimulation targets in MNI space: A subcortical atlas based on multimodal MRI, histology and structural connectivity. NeuroImage. 2018;170:271–282. 10.1016/j.neuroimage.2017.05.015

60. Chakravarty MM, Bertrand G, Hodge CP, Sadikot AF, Collins DL. The creation of a brain atlas for image guided neurosurgery using serial histological data. NeuroImage. 2006;30(2):359–376. doi:10.1016/j.neuroimage.2005.09.041

61. Treu S, Strange B, Oxenford S, et al. Deep brain stimulation: Imaging on a group level. NeuroImage. 2020;219:117018. 10.1016/j.neuroimage.2020.117018

62. Glasser MF, Sotiropoulos SN, Wilson JA, et al. The minimal preprocessing pipelines for the Human Connectome Project. NeuroImage. 2013;80:105–124. doi:10.1016/j.neuroimage.2013.04.127

63. Glasser MF, Smith SM, Marcus DS, et al. The Human Connectome Project’s neuroimaging approach. Nature Neuroscience. 2016;19(9):1175–1187. doi:10.1038/nn.4361

64. Jenkinson M, Bannister P, Brady M, Smith S. Improved Optimization for the Robust and Accurate Linear Registration and Motion Correction of Brain Images. NeuroImage. 2002;17(2):825–841. doi:10.1006/nimg.2002.1132

65. Behrens TEJ, Woolrich MW, Jenkinson M, et al. Characterization and propagation of uncertainty in diffusion-weighted MR imaging. Magnetic Resonance in Medicine. 2003;50(5):1077–1088. doi:10.1002/mrm.10609

66. Behrens TEJ, Berg HJ, Jbabdi S, Rushworth MFS, Woolrich MW. Probabilistic diffusion tractography with multiple fibre orientations: What can we gain? NeuroImage. 2007;34(1):144–155. doi:10.1016/j.neuroimage.2006.09.018

67. Elam JS, Glasser MF, Harms MP, et al. The Human Connectome Project: A retrospective. NeuroImage. 2021;244:118543. doi:10.1016/j.neuroimage.2021.118543

68. Remore LG, Rifi Z, Nariai H, et al. Structural connections of the centromedian nucleus of thalamus and their relevance for neuromodulation in generalized drug-resistant epilepsy: insight from a tractography study. Ther Adv Neurol Disord. 2023;16:17562864231202064. doi:10.1177/17562864231202064

69. Tsolaki E, Espinoza R, Pouratian N. Using probabilistic tractography to target the subcallosal cingulate cortex in patients with treatment resistant depression. Psychiatry Research: Neuroimaging. 2017;261:72–74. doi:10.1016/j.pscychresns.2017.01.006

70. Tsolaki E, Sheth SA, Pouratian N. Variability of white matter anatomy in the subcallosal cingulate area. Human Brain Mapping. 2021;42(7):2005–2017. doi:10.1002/hbm.25341

71. Shulman LM, Gruber-Baldini AL, Anderson KE, Fishman PS, Reich SG, Weiner WJ. The Clinically Important Difference on the Unified Parkinson’s Disease Rating Scale. Archives of Neurology. 2010;67(1):64–70. doi:10.1001/archneurol.2009.295

72. De Waele S, Hallemans A, Maréchal E, Cras P, Crosiers D. Gait initiation in Parkinson’s disease: comparison of timing and displacement during anticipatory postural adjustments as a function of motor severity and apathy in a large cohort. Heliyon. 2024;10(1):e23740. doi:10.1016/j.heliyon.2023.e23740

73. Palmisano C, Brandt G, Vissani M, et al. Gait Initiation in Parkinson’s Disease: Impact of Dopamine Depletion and Initial Stance Condition. Frontiers in Bioengineering and Biotechnology. 2020;8:137. doi:10.3389/fbioe.2020.00137

74. Bishop MD, Brunt D, Kukulka C, Tillman MD, Pathare N. Braking impulse and muscle activation during unplanned gait termination in human subjects with parkinsonism. Neuroscience Letters. 2003;348(2):89–92. doi:10.1016/S0304-3940(03)00738-9

75. Lu C, Twedell E, Elbasher R, McCabe M, MacKinnon CD, Cooper SE. Avoiding Virtual Obstacles During Treadmill Gait in Parkinson’s Disease. Frontiers in Aging Neuroscience. 2019;Volume 11-2019. https://www.frontiersin.org/journals/aging-neuroscience/articles/10.3389/fnagi.2019.00076

76. Caetano MJD, Lord SR, Allen NE, et al. Stepping reaction time and gait adaptability are significantly impaired in people with Parkinson’s disease: Implications for fall risk. Parkinsonism & Related Disorders. 2018;47:32–38. doi:10.1016/j.parkreldis.2017.11.340

77. Muro-de-la-Herran A, Garcia-Zapirain B, Mendez-Zorrilla A. Gait Analysis Methods: An Overview of Wearable and Non-Wearable Systems, Highlighting Clinical Applications. Sensors. 2014;14(2):3362–3394. doi:10.3390/s140203362

78. Hulleck AA, Menoth Mohan D, Abdallah N, El Rich M, Khalaf K. Present and future of gait assessment in clinical practice: Towards the application of novel trends and technologies. Frontiers in Medical Technology. 2022;Volume 4-2022. https://www.frontiersin.org/journals/medical-technology/articles/10.3389/fmedt.2022.901331

79. Gardoni A, Agosta F, Sarasso E, et al. Cerebellar alterations in Parkinson’s disease with postural instability and gait disorders. Journal of Neurology. 2023;270(3):1735–1744. doi:10.1007/s00415-022-11531-y

80. Madetko-Alster N, Alster P, Lamoš M, et al. The role of the somatosensory cortex in self-paced movement impairment in Parkinson’s disease. Clinical Neurophysiology. 2025;171:11–17. doi:10.1016/j.clinph.2025.01.001

81. Peterson DS, Pickett KA, Duncan RP, Perlmutter JS, Earhart GM. Brain activity during complex imagined gait tasks in Parkinson disease. Clinical Neurophysiology. 2014;125(5):995–1005. doi:10.1016/j.clinph.2013.10.008

82. Ren Q, Zhao S, Yu R, et al. Thalamic-limbic circuit dysfunction and white matter topological alteration in Parkinson’s disease are correlated with gait disturbance. Frontiers in Aging Neuroscience. 2024;Volume 16-2024. doi:10.3389/fnagi.2024.1426754

83. Hinton DC, Thiel A, Soucy JP, Bouyer L, Paquette C. Adjusting gait step-by-step: Brain activation during split-belt treadmill walking. NeuroImage. 2019;202(May):116095. doi:10.1016/J.NEUROIMAGE.2019.116095

84. Nachev P, Kennard C, Husain M. Functional role of the supplementary and pre-supplementary motor areas. Nature Reviews Neuroscience. 2008;9(11):856–869. doi:10.1038/nrn2478

85. Li H, Liu N, Li Y, Weidner R, Fink GR, Chen Q. The Simon Effect Based on Allocentric and Egocentric Reference Frame: Common and Specific Neural Correlates. Scientific Reports. 2019;9(1):13727. doi:10.1038/s41598-019-49990-5

86. la Fougère C, Zwergal A, Rominger A, et al. Real versus imagined locomotion: a [18F]-FDG PET-fMRI comparison. NeuroImage. 2010;50(4):1589–1598. doi:10.1016/J.NEUROIMAGE.2009.12.060

87. Obeso I, Loayza FR, González-Redondo R, et al. The causal role of the subthalamic nucleus in the inhibitory network. Annals of the New York Academy of Sciences. 2024;1538(1):117–128. doi:10.1111/nyas.15193

88. Swann N, Poizner H, Houser M, et al. Deep Brain Stimulation of the Subthalamic Nucleus Alters the Cortical Profile of Response Inhibition in the Beta Frequency Band: A Scalp EEG Study in Parkinson’s Disease. J Neurosci. 2011;31(15):5721. doi:10.1523/JNEUROSCI.6135-10.2011

89. Ballanger B, van Eimeren T, Moro E, et al. Stimulation of the subthalamic nucleus and impulsivity: Release your horses. Annals of Neurology. 2009;66(6):817–824. doi:10.1002/ana.21795

90. Cohen RG, Klein KA, Nomura M, et al. Inhibition, Executive Function, and Freezing of Gait. Journal of Parkinson’s Disease. 2014;4(1):111–122. doi:10.3233/JPD-130221

91. Shine JM, Handojoseno AMA, Nguyen TN, et al. Abnormal patterns of theta frequency oscillations during the temporal evolution of freezing of gait in Parkinson’s disease. Clinical Neurophysiology. 2014;125(3):569–576. doi:10.1016/j.clinph.2013.09.006

92. Mitchell T, Potvin-Desrochers A, Lafontaine AL, Monchi O, Thiel A, Paquette C. Cerebral Metabolic Changes Related to Freezing of Gait in Parkinson Disease. J Nucl Med. 2019;60(5):671. doi:10.2967/jnumed.118.218248

93. Nambu A, Tokuno H, Takada M. Functional significance of the cortico-subthalamo-pallidal “hyperdirect” pathway. Neuroscience research. 2002;43(2):111–117. doi:10.1016/S0168-0102(02)00027-5

94. Oswal A, Cao C, Yeh CH, et al. Neural signatures of hyperdirect pathway activity in Parkinson’s disease. Nature Communications. 2021;12(1):5185. doi:10.1038/s41467-021-25366-0

95. Chen W, de Hemptinne C, Miller AM, et al. Prefrontal-Subthalamic Hyperdirect Pathway Modulates Movement Inhibition in Humans. Neuron. 2020;106(4):579–588.e3. doi:10.1016/j.neuron.2020.02.012

96. Favre E, Ballanger B, Thobois S, Broussolle E, Boulinguez P. Deep Brain Stimulation of the Subthalamic Nucleus, but not Dopaminergic Medication, Improves Proactive Inhibitory Control of Movement Initiation in Parkinson’s Disease. Neurotherapeutics. 2013;10(1):154–167. doi:10.1007/s13311-012-0166-1

