## Supplementary for "Subthalamic DBS Engages Right-lateralized Frontal Control to Improve Gait Adaptation in Parkinson’s"

Supplementary table 1: Subjects' clinical characteristics in stim-off and stim-on conditions based on MDS-UPDRS-III

| Subject | Stim-off | | | Stim-on | | | DBS-induced change | | |
|---|---|---|---|---|---|---|---|---|---|
| **Items** | **Total** | **PIGD** | **Axial** | **Total** | **PIGD** | **Axial** | **Total** | **PIGD** | **Axial** |
| BOS01 | 39 | 1 | 3 | 21 | 1 | 3 | -18 | 0 | 0 |
| BOS03 | 34 | 5 | 8 | 19 | 1 | 3 | -15 | -4 | -5 |
| BOS04 | 58 | 2 | 4 | 24 | 1 | 3 | -34 | -1 | -1 |
| PER02 | 47 | 4 | 7 | 30 | 2 | 3 | -17 | -2 | -4 |
| PER11 | 45 | 2 | 4 | 20 | 2 | 3 | -25 | 0 | -1 |
| PER17 | 36 | 1 | 2 | 18 | 0 | 1 | -18 | -1 | -1 |
| PER18 | 17 | 0 | 1 | 5 | 0 | 0 | -12 | 0 | -1 |
| PER19 | 78 | 5 | 10 | 45 | 3 | 7 | -33 | -2 | -3 |
| PER20 | 44 | 6 | 10 | 21 | 4 | 7 | -23 | -2 | -3 |
| PER22 | 56 | 5 | 9 | 27 | 3 | 6 | -29 | -2 | -3 |
| PER26 | 33 | 3 | 5 | 22 | 2 | 4 | -11 | -1 | -1 |
| PER33 | 47 | 2 | 4 | 15 | 1 | 2 | -32 | -1 | -2 |
| **Mean** | 44.3 | 3.0 | 5.6 | 22.3 | 1.7 | 3.5 | -22.1 | -1.3 | -2.1 |
| **SD** | 15.4 | 2.0 | 3.1 | 9.5 | 1.2 | 2.2 | 8.5 | 1.2 | 1.5 |
| **P value** | Paired-samples student t test | | | | | | <0.001 | 0.002 | <0.001 |

Supplementary table 2: Stimulation parameters of the subject cohort

| Subject | Left Amplitude [mA] | Right Amplitude [mA] | AI Stim | Frequency [Hz] | Pulse width [µs] |
|---|---|---|---|---|---|
| BOS01 | 2.60 | 4.60 | -0.28 | 185 | 60 |
| BOS03 | 3.20 | 2.40 | 0.14 | 130 | 40 |
| BOS04 | 4.20 | 4.60 | -0.05 | 179 | 55 |
| PER02 | 3.30 | 2.90 | 0.06 | 180 | 60 |
| PER11 | 3.30 | 4.60 | -0.16 | 125 | 60 |
| PER17 | 4.10 | 2.90 | 0.17 | 180 | 60 |
| PER18 | 3.50 | 4.50 | -0.13 | 125 | 60 |
| PER19 | 4.30 | 3.90 | 0.05 | 125 | 60 |
| PER20 | 3.30 | 3.00 | 0.05 | 125 | 60 |
| PER22 | 3.00 | 3.00 | 0.00 | 125 | 60 |
| PER26 | 5.00 | 3.00 | 0.25 | 125 | 60 |
| PER33 | 2.20 | 3.10 | -0.17 | 180 | 60 |

*AI Stim: Positive values indicate higher stimulation intensity on the left hemisphere and vice versa.*

Supplementary table 3: Principal component analysis for gait performance parameters during obstacle avoidance

| | Stim-off | | Stim-on | |
|---|---|---|---|---|
| KMO measure of sampling adequacy | 0.611 | | 0.788 | |
| Bartlett's test of sphericity | P<0.001 | | P<0.001 | |
| **Components*** | **1 (Gait adaptation)** | **2 (Min. SVAD)** | **1 (Gait adaptation)** | **2 (Min. SVAD)** |
| Initial Eigenvalue | 4.3 | 1.4 | 4.8 | 0.9 |
| Eigenvalue after rotation | 4.3 | 1.4 | 3.9 | 1.8 |
| % of variance | 71.0 | 23.6 | 64.6 | 30.2 |
| **Rotated features loading†** | | | | |
| AW duration | -0.958 | - | -0.833 | - |
| Deceleration | -0.948 | - | -0.988 | - |
| Acceleration | 0.973 | - | 0.934 | - |
| V. VA start | 0.978 | - | 0.910 | - |
| V. Minimum | - | - | - | - |
| Min. SVAD | - | 0.975 | - | 0.972 |

** The PCA was ran with two as a fixed number of extracted factors, using Varimax methods for factor rotation with Kaise; † Features with an absolute loading value <0.70 were suppressed; KMO: Kaiser-Meyer-Olkin (KMO) test for sampling adequacy; AW: Adaptation window; V.: Velocity; Min. SVAD: Minimum subject-virtual agent distance.*

Supplementary table 4: The demographic characteristics and kinematic features of healthy subjects comparable in age and gender to the subjects with PD

| **Subject** | **Gender** | **Age [year]** | **AW Duration [s]** | **Deceleration [m/s$^2$]** | **Acceleration [m/s$^2$]** | **V. VA start [m/s]** |
|---|---|---|---|---|---|---|
| OHS01 | F | 60-65 | 1.04 | -0.71 | 1.20 | 1.34 |
| OHS02 | F | 60-65 | 1.31 | -0.59 | 1.29 | 1.12 |
| OHS03 | M | 56-60 | 1.11 | -0.82 | 1.54 | 1.50 |
| OHS05 | M | 66-70 | 1.67 | -0.60 | 1.11 | 1.22 |
| OHS06 | M | 56-60 | 1.47 | -0.61 | 1.01 | 1.11 |
| OHS07 | M | 66-70 | 1.48 | -0.67 | 1.31 | 1.15 |
| OHS08 | M | 70-75 | 1.34 | -0.58 | 1.32 | 1.23 |
| OHS10 | M | 60-65 | 1.76 | -0.57 | 1.06 | 1.06 |
| **Mean** | 6/2 | 63.3 | 1.40 | -0.64 | 1.23 | 1.22 |
| **SD** | M/F | 5.8 | 0.25 | 0.08 | 0.17 | 0.14 |

*AW: Adaptation window; V.: Velocity.*

Supplementary table 5: The kinematic features significantly contributing to the first principal component in Walk-off and Walk-on conditions, and their percentual change across conditions

| | Walk-off | | | | Walk-on | | | | Improvement due to DBS | | | |
|---|---|---|---|---|---|---|---|---|---|---|---|---|
| Subject | AW duration [s] | Deceleration [m/s$^2$] | Acceleration [m/s$^2$] | V. at VA start [m/s] | AW duration [s] | Deceleration [m/s$^2$] | Acceleration [m/s$^2$] | V. at VA start [m/s] | AW duration [s] | Deceleration [m/s$^2$] | Acceleration [m/s$^2$] | V. at VA start [m/s] |
| BOS01 | 2.61 | -0.32 | 0.53 | 0.76 | 2.71 | -0.34 | 0.63 | 0.76 | 3.6% | 7.0% | 18.7% | -5.0% |
| BOS03 | 2.10 | -0.35 | 0.66 | 0.79 | 1.81 | -0.63 | 1.00 | 1.01 | -13.6% | 78.2% | 53.0% | 26.3% |
| BOS04 | 2.41 | -0.35 | 0.59 | 0.68 | 2.16 | -0.42 | 0.81 | 0.77 | -10.2% | 21.2% | 37.0% | 10.0% |
| PER02 | 3.84 | -0.17 | 0.34 | 0.48 | 3.29 | -0.20 | 0.40 | 0.49 | -14.3% | 21.5% | 17.8% | -2.0% |
| PER11 | 2.17 | -0.62 | 0.65 | 0.94 | 1.91 | -0.56 | 0.93 | 0.94 | -11.7% | -9.6% | 42.8% | 4.4% |
| PER17 | 1.45 | -0.41 | 0.75 | 1.01 | 1.41 | -0.46 | 1.17 | 1.06 | -3.0% | 12.1% | 56.9% | 6.0% |
| PER18 | 2.10 | -0.50 | 0.77 | 0.77 | 2.13 | -0.67 | 1.12 | 0.91 | 1.3% | 33.6% | 44.7% | 13.8% |
| PER19 | 8.53 | -0.02 | 0.08 | 0.14 | 3.28 | -0.16 | 0.36 | 0.46 | -61.6% | 706.8% | 350.4% | 360.0% |
| PER20 | 7.65 | -0.05 | 0.09 | 0.23 | 4.96 | -0.10 | 0.22 | 0.32 | -35.0% | 78.0% | 143.0% | 60.0% |
| PER22 | 4.40 | -0.11 | 0.19 | 0.38 | 3.22 | -0.15 | 0.38 | 0.50 | -26.8% | 43.5% | 94.8% | 25.0% |
| PER33 | 2.52 | -0.40 | 0.72 | 0.70 | 2.00 | -0.52 | 0.84 | 0.87 | -20.7% | 27.8% | 15.5% | 24.3% |
| **Mean** | 3.62 | -0.30 | 0.49 | 0.63 | 2.63 | -0.38 | 0.71 | 0.74 | -17.5% | 92.7% | 79.5% | 47.5% |
| **SD** | 2.37 | 0.19 | 0.27 | 0.28 | 1.02 | 0.20 | 0.34 | 0.25 | 18.6% | 205.5% | 97.4% | 105.2% |
| P value | Paired-samples Wilcoxon rank test for the difference between Walk-off and Walk-on | | | | | | | | 0.010 | 0.016 | 0.003 | 0.006 |

*AW: Adaptation window; V.: Velocity; VA: Virtual agent.*

Supplementary table 6: Correlations between the kinematic features and clinical assessments by MDS-UPDRS-III score and sub-scores

| | **Stim-off** | | | | | | **Stim-on** | | | | | |
|---|---|---|---|---|---|---|---|---|---|---|---|---|
| Pearson r | Total | PIGD | Axial | Rigidity | Tremor | Brady. | Total | PIGD | Axial | Rigidity | Tremor | Brady. |
| AW duration‡ | 0.64 | 0.73 | **0.80†** | 0.60 | 0.24 | 0.50 | 0.41 | **0.86†** | **0.79†** | 0.01 | 0.09 | 0.27 |
| Deceleration‡ | 0.61 | **0.78†** | **0.83†** | 0.46 | 0.28 | 0.43 | 0.72 | **0.81†** | **0.83†** | -0.11 | 0.31 | 0.63 |
| Acceleration‡ | -0.66 | **-0.80†** | **-0.87†** | -0.48 | -0.20 | -0.59 | -0.67 | **-0.89†** | **-0.87†** | 0.06 | -0.26 | -0.56 |
| V. VA start‡ | -0.64 | **-0.76†** | **-0.83†** | -0.48 | -0.36 | -0.41 | -0.61 | **-0.87†** | **-0.83†** | -0.02 | -0.30 | -0.46 |
| Composite score | -0.66 | **-0.79†** | **-0.86†** | -0.53 | -0.28 | -0.50 | -0.63 | **-0.88†** | **-0.85†** | 0.04 | -0.25 | -0.50 |

*Cell numbers represent the Pearson r correlation coefficient between the kinematic features and the sub-scores of MDS-UPDRS-III; ‡ These variables are part of PC1; † Significant correlations with a P < 0.01; Brady.: Bradykinesia; Min. SVAD: Minimum subject-virtual agent distance; AW: Adaptation window; PIGD: Postural instability and gait disorder; V.: Velocity; Composite score: The combined kinematic performance measure that is defined as the average Z-values of all features within PC1.*

Supplementary table 7: Coordinates and significance levels of the FDG-PET contrast analysis

| Cluster | | | Peak | | | | Coordinates | | | |
|---|---|---|---|---|---|---|---|---|---|---|
| **equivk** | **p(unc)** | **p(FWE-corr)** | **F** | **equivZ** | **p(unc)** | **p(FWE-corr)** | **X*** | **Y*** | **Z*** | **Volume of interest** |
| **F-test across the three FDG-PET scans (P<0.001)** | | | | | | | | | | |
| 7095 | <0.001 | <0.001 | 72.42 | 6.25 | <0.001 | <0.001 | **0** | **-50** | **-10** | **Vermis** |
| | | | 63.22 | 6.04 | <0.001 | <0.001 | 8 | -50 | -18 | Right Cerebellum |
| | | | 52.38 | 5.76 | <0.001 | 0.001 | 2 | -64 | -26 | Vermis |
| 4195 | <0.001 | <0.001 | 44.89 | 5.52 | <0.001 | 0.002 | **-2** | **-30** | **66** | **Left Paracentral Lobule** |
| | | | 42.30 | 5.43 | <0.001 | 0.003 | 6 | -32 | 68 | Right Paracentral Lobule |
| | | | 34.07 | 5.09 | <0.001 | 0.013 | -4 | -42 | 52 | Left Mid Cingulum |
| 1099 | <0.001 | <0.001 | 25.46 | 4.62 | <0.001 | 0.085 | **2** | **40** | **-8** | **Right Orb Med Frontal Gyrus** |
| | | | 21.44 | 4.35 | <0.001 | 0.220 | 2 | 18 | -6 | Right Olfactory Gyrus |
| | | | 17.93 | 4.06 | <0.001 | 0.488 | 4 | 58 | 0 | Right Orb Med Frontal Gyrus |
| **Hypermetabolism in Walk-off vs. Resting (P=0.002)** | | | | | | | | | | |
| 5220 | <0.001 | <0.001 | 10.64 | 6.26 | <0.001 | <0.001 | **8** | **-52** | **-18** | **Right Cerebellum** |
| | | | 10.44 | 6.20 | <0.001 | <0.001 | 0 | -50 | -10 | Vermis |
| | | | 9.08 | 5.79 | <0.001 | <0.001 | 16 | -42 | -20 | Right Cerebellum |
| 4010 | <0.001 | <0.001 | 7.25 | 5.13 | <0.001 | 0.008 | **6** | **-32** | **68** | **Right Paracentral Lobule** |
| | | | 6.93 | 5.00 | <0.001 | 0.014 | -2 | -30 | 66 | Left Paracentral Lobule |
| | | | 6.62 | 4.86 | <0.001 | 0.025 | -6 | -52 | 56 | Left Precuneus |
| **Hypometabolism in Walk-off vs. Resting (P=0.057)** | | | | | | | | | | |
| 2412 | <0.001 | <0.001 | 6.95 | 5.00 | <0.001 | 0.014 | **2** | **40** | **-8** | **Right Orb Med Frontal Gyrus** |
| | | | 5.98 | 4.56 | <0.001 | 0.079 | 4 | 58 | 0 | Right Orb Med Frontal Gyrus |
| | | | 5.77 | 4.45 | <0.001 | 0.116 | **-14** | 46 | -16 | Left Orb Med Frontal Gyrus |
| **Hypermetabolism in Walk-on vs. Walk-off (P=0.006)** | | | | | | | | | | |
| 217 | 0.012 | 0.086 | 5.33 | 4.22 | <0.001 | 0.243 | **-14** | **-10** | **-1** | **Left Thalamus** |
| 213 | 0.013 | 0.091 | 5.18 | 4.15 | <0.001 | 0.306 | **-12** | **20** | **52** | **Left Sup Frontal Gyrus** |
| | | | 4.31 | 3.63 | <0.001 | 0.839 | -12 | 32 | 44 | Left Sup Frontal Gyrus |
| | | | 4.22 | 3.57 | <0.001 | 0.884 | -14 | 8 | 56 | Left Sup Frontal Gyrus |
| **Hypometabolism in Walk-on vs. Walk-off (P=0.411)** | | | | | | | | | | |
| 161 | 0.027 | 0.180 | 4.97 | 4.03 | <0.001 | 0.418 | **12** | **-56** | **-14** | **Right Cerebellum** |
| | | | 4.40 | 3.69 | <0.001 | 0.788 | 12 | -66 | -16 | Right Cerebellum |
| | | | 3.90 | 3.36 | <0.001 | 0.977 | 22 | -52 | -22 | Right Cerebellum |

** The coordinates are based on the anterior commissure as the origine (0,0,0) and the RAS+ convention in the MNI space.*

Supplementary table 8: Stepwise linear regression demonstrating the independent contributions of asymmetry in clinical impairment, delivered stimulation power, and metabolic SFG activity towards obstacle avoidance improvement

| Variables | ORs | P value |
|---|---|---|
| **Dependent variable: DBS-related composite score change** | | |
| Step 1 | F=2.096, $R^2$=0.248 | P=0.189 |
| AI Clinic | 0.069 [-1.503, 1.801] | 0.837 |
| AI Stim | -0.117 [-2.805, 2.240] | 0.799 |
| AI PET change | -0.791 [-0.503, 0.083] | 0.135 |
| Step 2 | F=3.545, $R^2$=0.337 | P=0.079 |
| AI Stim | -0.076 [-2.271, 1.901] | 0.843 |
| AI PET change | -0.739 [-0.424, 0.033] | 0.084 |
| Step 3 | F=7.888, $R^2$=0.408 | P=0.020 |
| AI PET change | -0.683 [-0.327, -0.035] | 0.020 |
| **Dependent variable: DBS-related AI PET change** | | |
| Step 1 | F=7.625, $R^2$=0.570 | P=0.014 |
| AI Clinic | 0.360 [-0.969, 6.865] | 0.121 |
| AI Stim | -0.738 [-11.103, -2.370] | 0.007 |
| Step 2 | F=10.001, $R^2$=0.474 | P=0.012 |
| AI Stim | -0.725 [-11.354, -1.884] | 0.012 |
| **Dependent variable: AI stim** | | |
| Step 1 | F=0.011, $R^2$=-0.110 | P=0.917 |
| AI Clinic | 0.036 [-0.644, 0.708] | 0.917 |

*The regression used backward methods; $R^2$ values are adjusted for the number of included variables.*

Supplementary figure 1: Leads and active contacts localization with respect to the targeted subthalamic nuclei. The first row shows a lateral and superior view of the lead localization with respect to the bilateral subthalamic nuclei in orange; The lower row shows lateral views of the active contacts placement in relation to the targeted subthalamic nuclei; All figures show the subthalamic nuclei and the placement within the normative MNI space. For the visualization of this figure, participants were stratified into three groups according to their level of DBS-related composite gait-adaptation score change (low, moderate, or high), using cluster analysis with Ward's linkage method.

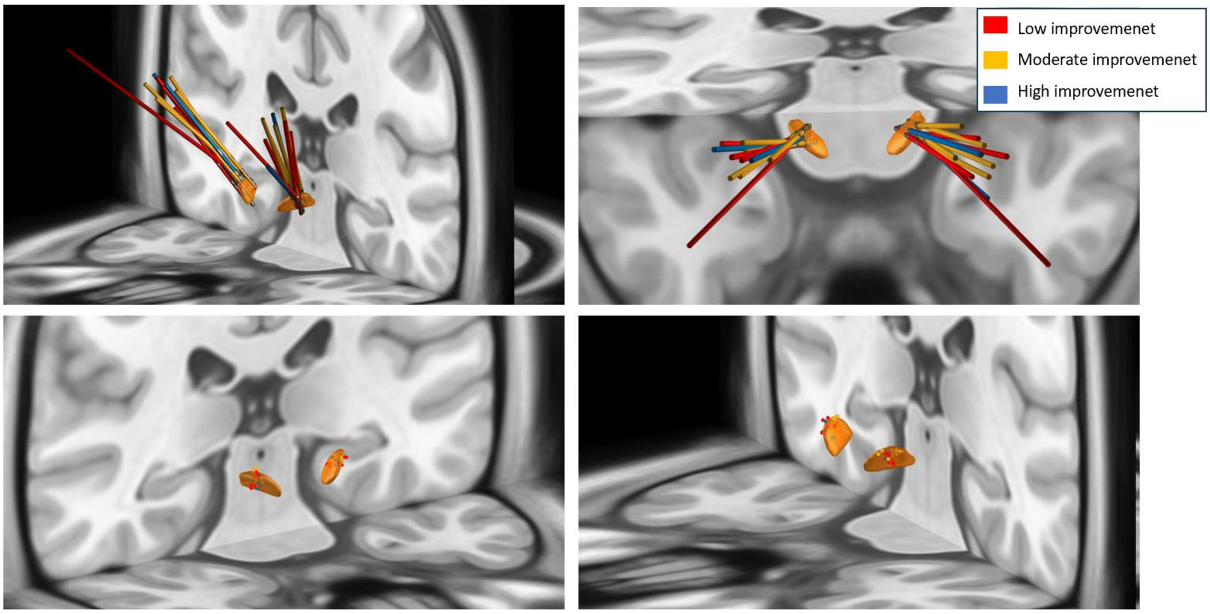

Supplementary figure 2: A visualization of the normative structural connectivity between the estimated VTAs and the activated region of the SFG. Left panel: structural connections from STN to SFG for the three patients high improvement (blue), moderate improvement (yellow), and low improvement (red) groups. Right panel: Structural connection from STN to SFG without mask-constriction (ocher) and with the thalamus as way-point mask (yellow). The latter connection has been calculated by forcing the output fibers from the STN to travel through the thalamus (blue); thus, the resulting tract is thinner and more direct. The connections calculated with FSL software have been non-linearly coregistered to the MNI152 NLIN 2009b space and visualized in Lead-DBS viewer with the 7 T MNI 2009b asymmetric T1 as background. Purple: a mask of the cluster of the SFG showing higher FDG uptake on stim-on; Orange: STN. For the visualization of this figure, participants were stratified into three groups according to their level of DBS-related composite gait-adaptation score change (low, moderate, or high), using cluster analysis with Ward's linkage method.

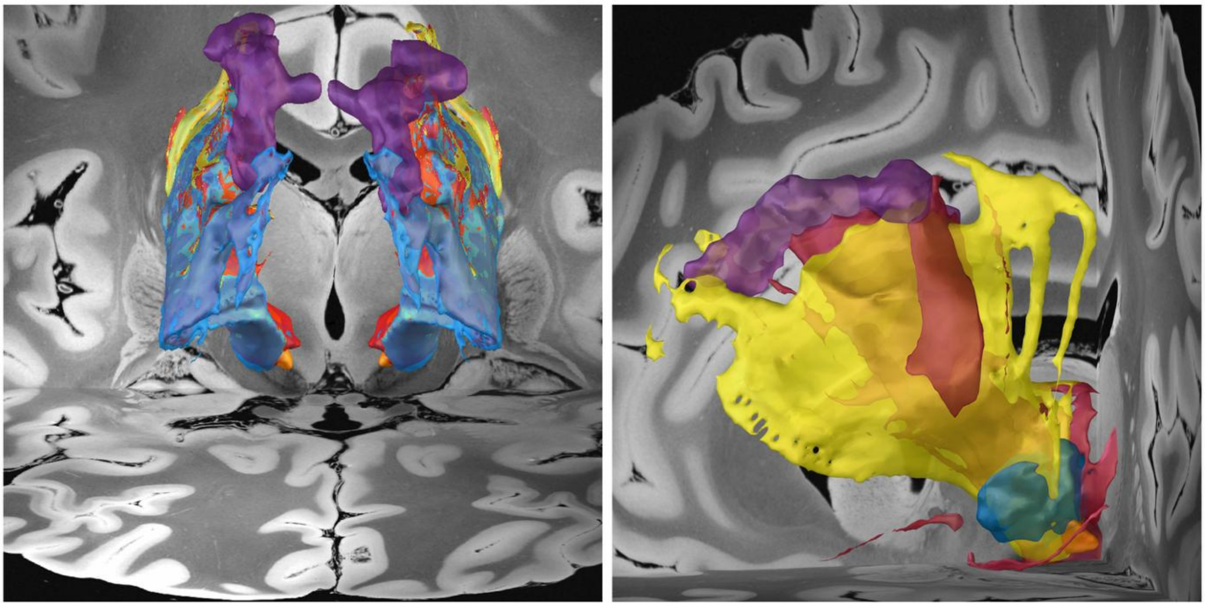

Supplementary figure 3: Correlation between DBS-related composite score change and the normalized intersection between the STN and the volume of tissue activated. All figures show the normalized intersection between the estimated volume of tissue activated and the full STN (upper raw) or the motor STN (lower raw); The left panels refer to the left hemisphere, the middle ones refer to the right hemisphere, and the right ones refer to the asymmetry indices between the left and right hemispheres.

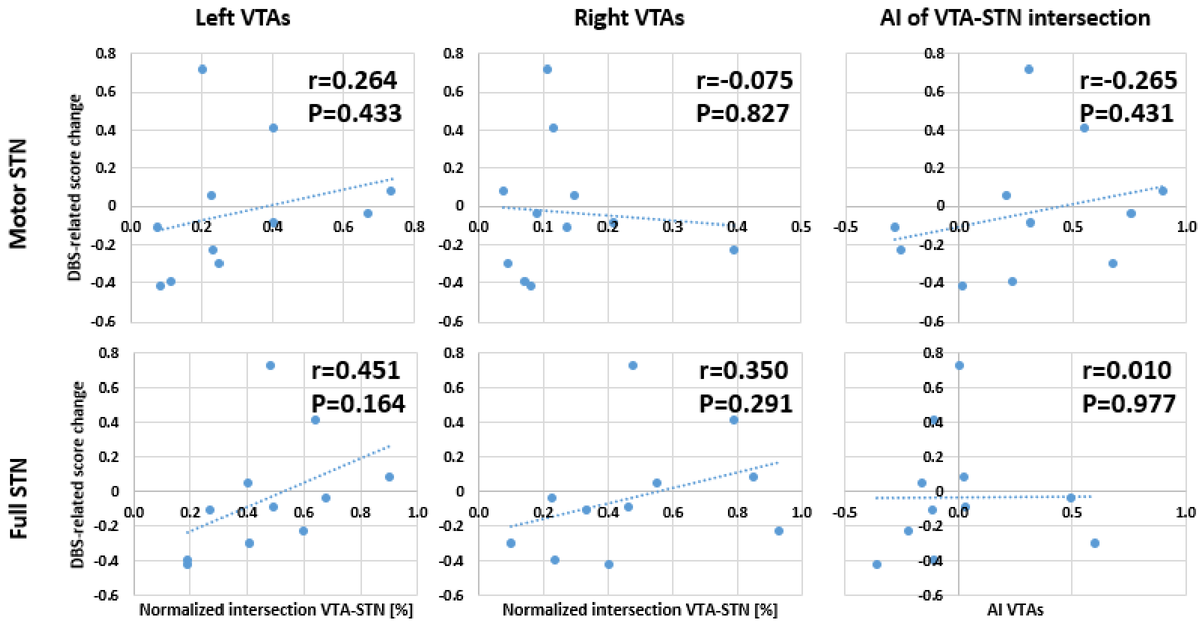

Supplementary figure 4: Correlation between the probabilistic connectivity measures and DBS-related gait adaptation improvement. The probability of connectivity for the left two panels was measured between the estimated VTAs and a mask of the SFG cluster showing significant increase of uptake in Walk-on with regards to Walk-off; The right panel shows a correlation between the DBS-related composite score improvement and the asymmetry index of the bilateral probability of connectivity; AI: asymmetry index; r: Pearson correlation coefficient.

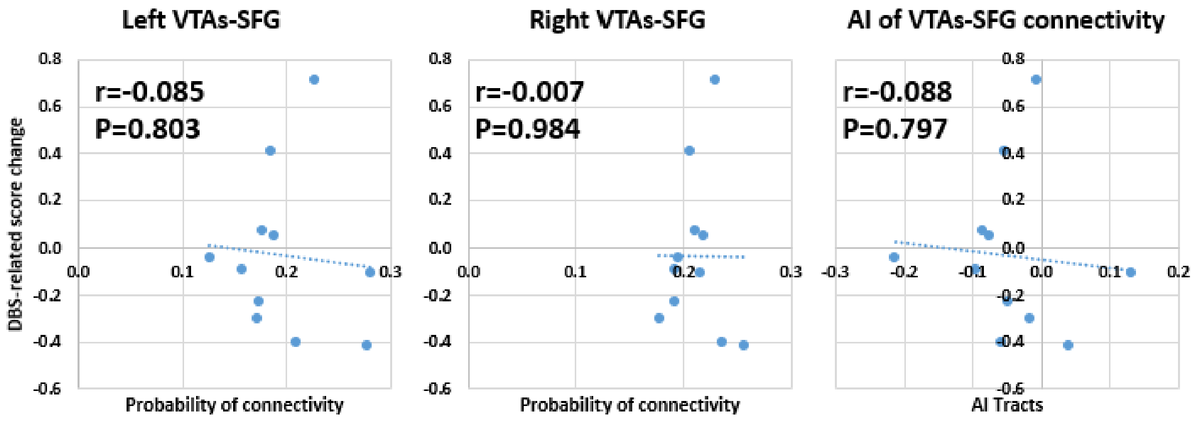

Supplementary table 9: Correlation between the asymmetry indices of clinical impairment, delivered stimulation power, VTAs-STN intersection, and VTAs-SFG probability of connectivity on one side and the metabolic activation asymmetry and composite gait-adaptation score change on the other side.

| | **AI PET** | | | **Composite gait-adaptation score** | | |
|---|---|---|---|---|---|---|
| | **Walk-off** | **Walk-on** | **DBS-change** | **Walk-off** | **Walk-on** | **DBS-change** |
| AI Clinic* | 0.537 | **0.644**† | 0.334 | -0.340 | -0.418 | -0.200 |
| AI Stim‡ | -0.020 | -0.228 | **-0.725**† | -0.292 | -0.131 | 0.460 |
| AI VTAs§ | -0.367 | -0.325 | -0.048 | 0.455 | 0.559 | 0.265 |
| AI Tracts | -0.066 | -0.041 | 0.132 | -0.067 | -0.100 | -0.088 |

** Higher values indicate more impairment on the right hemisphere/left hemibody as compared to the contralateral hemisphere/hemibody; † Statistically significant at α=0.05; ‡ Higher values demonstrate higher relative power on the left hemisphere electrode; § Calculated on the motor segment of the STN.*

Supplementary table 10: Kinematic gait features unrelated to the gait adaptation performance (i.e., steady-state velocity and peak turning velocity) in the Walk-off and Walk-on conditions

| | **Walk-off** | | **Walk-on** | | **DBS-related improvement** | |
|---|---|---|---|---|---|---|
| **Subjects with PD** | **Steady state velocity [m/s]** | **Peak turning velocity [rad/s]** | **Steady state velocity [m/s]** | **Peak turning velocity [rad/s]** | **Steady state velocity [m/s]** | **Peak turning velocity [rad/s]** |
| BOS01 | 0.68 | 68.19 | 0.71 | 89.38 | 4.0% | 31.1% |
| BOS03 | 0.74 | 71.54 | 0.96 | 114.42 | 29.7% | 59.9% |
| BOS04 | 0.62 | 84.46 | 0.72 | 117.74 | 14.7% | 39.4% |
| PER02 | 0.48 | 51.47 | 0.49 | 58.99 | 3.7% | 14.6% |
| PER11 | 0.87 | 75.04 | 0.97 | 120.61 | 12.0% | 60.7% |
| PER17 | 0.92 | 105.54 | 0.93 | 113.41 | 0.7% | 7.5% |
| PER18 | 0.74 | 106.15 | 0.97 | 153.73 | 32.1% | 44.8% |
| PER19 | 0.11 | - | 0.44 | - | 289.2% | - |
| PER20 | 0.22 | - | 0.38 | - | 71.3% | - |
| PER22 | 0.38 | 54.78 | 0.46 | 65.43 | 21.2% | 19.4% |
| PER33 | 0.73 | 84.26 | 0.79 | 97.42 | 8.0% | 15.6% |
| **Mean** | 0.59 | 77.94 | 0.71 | 103.46 | 44.2% | 32.6% |
| **SD** | 0.26 | 19.43 | 0.23 | 29.37 | 83.7% | 19.8% |
| **P value** | Paired-samples Wilcoxon rank test for the difference Walk-off vs. Walk-on | | | | 0.003 | 0.008 |

*Steady state velocity represents the average velocity in the 1s before the start of the virtual agent; Peak turning velocity represents the maximal angular velocity of the turns in between subsequent gait adaptation trials; two subjects with PD were unable to turn without assistance, which yielded only nine subjects with PD for the turning velocity analysis; DBS-related improvement represents the relative change of each variable from Walk-off to Walk-on condition.*

Supplementary figure 5: Pearson correlations between the asymmetry in the metabolic activity of the SFG and kinematic gait variables unrelated to obstacle avoidance (i.e., steady-state velocity and peak turning velocity). The x axis of each Walk-off and Walk-on panel depicts the Z value of the corresponding variable; AI PET SFG: asymmetry index of the FDG uptake in the SFG, where positive values demonstrate greater relative uptake in the left hemisphere; DBS-related AI change: change in the AI PET SFG in Walk-on with respect to Walk-off, where values are positive when DBS induces higher FDG uptake in the left SFG, and negative when it favors higher FDG uptake on the right SFG.

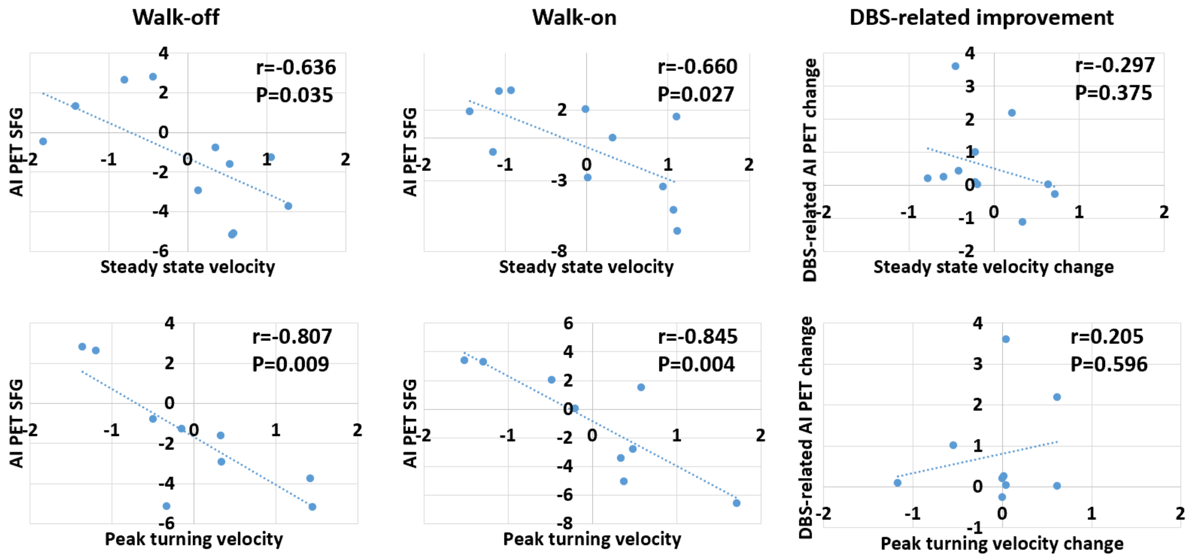
